# Hearing Loss and Loneliness in the Development of Impaired Cognition and Dementia in the All of Us Cohort: Prospective Analyses Using Electronic Health Records

**DOI:** 10.1101/2025.11.11.25340017

**Authors:** Shuai Yang, David A. Sbarra, Jingyue Wu, Yann C. Klimentidis

**Affiliations:** Department of Epidemiology and Biostatistics, Mel & Enid Zuckerman College of Public Health, University of Arizona, Tucson, AZ, USA; Department of Psychology, University of Arizona, Tucson, AZ, USA

**Keywords:** hearing loss, loneliness, dementia, impaired cognition, mediation, Cox models, All of Us

## Abstract

**Background:** Hearing loss (HL) and loneliness have each been linked to cognitive decline, yet the extent to which loneliness mediates the HL–cognition relationship remains uncertain. We examined prospective associations among HL, loneliness, and incident dementia and impaired cognition (IC), and tested whether loneliness partially mediates the effect of HL on these outcomes.

**Methods:** Using the NIH All of Us (AoU) Controlled Tier v8 dataset, we fit adjusted Cox proportional hazards models with distinct baselines for HL (first EHR visit) and loneliness (social determinants of health survey). Loneliness was measured with the UCLA Loneliness Scale–8 (ULS-8) and analyzed as the ULS-8 mean. We conducted four-way decomposition mediation analyses to partition total, direct, and indirect effects, allowing for exposure–mediator interaction. Outcomes (incident dementia and IC) were ascertained from EHRs after the relevant baseline exposure. Sensitivity analyses examined associations in racial/ethnic subgroups, and used time-varying HL in Cox models. Covariates included age, sex, race, education, income, smoking, and alcohol use.

**Findings:** The analytic sample comprised 317,020 adults (mean overall follow-up, 11.8 years); 50,807 (16.0%) had EHR-documented HL and 132,424 (41.8%) completed the loneliness survey. We observed 6,042 incident dementia and 21,699 incident IC events. HL was associated with higher risk of incident dementia (HR = 1.29, p < .001) and IC (HR = 1.65, p < .001). Loneliness (per 1-point increase in ULS-8 mean) was associated with higher incident dementia (HR = 1.41, 95% CI 1.12–1.75, p = .003) and IC (HR = 1.49, 95% CI 1.36–1.62, p < .001) risk. Mediation analyses indicated small indirect effects of HL via loneliness. The proportion mediated was approximately 2.4% for dementia (pm = 0.024, p = .090) and 2.0% for IC (pm = 0.020, p = .0004). Subgroup analyses suggested stronger HL–dementia associations among Black participants and stronger HL–IC associations among Asian and Black participants, while results from time-varying models were largely confirmatory.

**Interpretation:** In this large, diverse cohort, HL and loneliness were independently associated with incident dementia and IC. Loneliness mediated only a small fraction of the HL effect, suggesting that hearing rehabilitation and social connection supports may be complementary strategies for cognitive health, with potentially more benefits from addressing HL directly.

**Funding:** This study was supported in part by an AoU University of Arizona–Banner Health Driver Grant Program award to Drs. Klimentidis and Sbarra, which was funded in part by the National Institutes of Health Office of the Director through the University of Arizona-Banner AoU Researcher’s Collective (award OT2OD036485). The content is solely the responsibility of the authors and does not necessarily represent the official views of the AoU Research Program, the National Institutes of Health, or any other funder. The funders had no role in the study design, data access, data analysis and interpretation, the decision to submit the work for publication, or the preparation of the manuscript.

## INTRODUCTION

Dementia imposes profound personal and societal costs, and among modifiable midlife risks, hearing loss (HL) stands out as a leading and actionable target. The 2020 Lancet Commission on dementia prevention estimated that untreated midlife HL may account for about 8% of dementia cases worldwide—a greater proportion than other established risk factors like hypertension or obesity (1). Several mechanisms are proposed to explain how hearing impairment might contribute to cognitive decline and dementia, including increased cognitive load (i.e., the brain needs to work harder to process degraded auditory input, diverting resources from memory and thinking), accelerated brain atrophy due to reduced sensory stimulation, and social disengagement (2–5). In particular, HL may contribute to social isolation, as difficulty hearing can prompt withdrawal from conversations and activities, reducing cognitive stimulation and resilience. Consistent with this pathway, observational studies show that HL is associated with higher odds of loneliness and social isolation in older adults (5). Population-based studies further support this association: greater HL is linked to higher odds of social isolation in older women (6), and in a nationally representative U.S. cohort, hearing impairment was associated with a 28% greater odds of social isolation over eight years (7). Relatedly, Medicare beneficiaries with functional HL had higher odds of limited social engagement (8). Together, these findings suggest that social disconnection is a plausible pathway linking HL with future cognitive decline and perhaps overall brain health.

Loneliness is widely recognized as an important psychosocial factor impacting cognitive health (9,10). A recent meta-analysis of longitudinal studies (*N* > 600,000) found that feeling lonely is associated with about a 31% higher risk of developing dementia (11). Loneliness is also linked to faster cognitive decline and broader adverse health outcomes(11–15). Notably, loneliness appears to influence dementia risk independently of objective social isolation (16), suggesting that the emotional experience of disconnection may have a unique impact on the brain.

There is a plausible interplay between HL and loneliness in the pathogenesis of dementia. Because hearing impairment often reduces social engagement, it may precipitate or exacerbate loneliness in some individuals. Loneliness, in turn, could be one pathway through which HL influences cognitive decline—a hypothesis consistent with a “cascade” model of dementia risk where sensory deficits lead to social isolation and subsequently to cognitive impairment (17). Empirical evidence of this mediation pathway, however, is limited. Prior studies show that HL is associated with increased loneliness and social isolation (5–8) and that both HL and loneliness independently elevate dementia risk (3,11,16,18); but whether, and to what extent, loneliness explains the HL–dementia link remains unclear. A recent systematic review found insufficient evidence that social isolation significantly mediates the association between HL and cognitive decline (18), highlighting the need for direct analyses of this question in large longitudinal datasets.

The present study leverages data from the National Institutes of Health All of Us (AoU) Research Program, a large, nationally diverse U.S. cohort, to investigate the interrelationships between HL, loneliness, and dementia risk. This large and diverse cohort—over 80% of participants are from groups historically underrepresented in biomedical research (19)—provides an opportunity to examine interrelationships among HL, loneliness, and cognitive outcomes, and to assess how these associations vary across race/ethnicity and socioeconomic subgroups. In addition, linkage to electronic health records (EHRs) for a large proportion of participants allows for more precise temporal ordering of exposures and outcomes, enhancing the validity of causal inferences (19). Although our work was not preregistered and should thus be considered exploratory in nature, we were guided by several specific aims designed to examine whether: (1) HL, diagnosed prior to the loneliness survey, is associated with greater future loneliness; (2) HL and loneliness independently predict the development of dementia and of impaired cognition (IC); and (3) loneliness mediates the effect of HL on these cognitive outcomes. By examining these questions in a large, diverse EHR-based cohort, this study aims to clarify the role of HL and loneliness in late-life cognitive health and to inform potential interventions (e.g., hearing rehabilitation and social engagement strategies) for dementia prevention.

## METHODS

### Study Design and Sample

We conducted an observational cohort study using data (Controlled Tier Dataset v8) from the AoU Research Program, a nationwide U.S. research cohort launched in 2018 by the NIH. AoU is enrolling at least one million adult participants from diverse communities across the United States to advance precision medicine (19). Participants contribute data through linked EHRs, survey questionnaires, physical measurements, and biospecimen samples. The AoU cohort is notable for its diversity: more than 80% of participants are from groups historically underrepresented in biomedical research, and the cohort includes a wide range of ages, socio-economic backgrounds, and racial/ethnic identities (19).

For this analysis, we included AoU participants from the v8 data release who had either an ascertainable baseline HL status from the EHR or a completed baseline loneliness survey and who had subsequent EHR follow-up for cognitive outcomes. We distinguish two anchors: the Hearing-Loss baseline (HL-baseline), defined as the first EHR visit (start of retrospective ascertainment for HL), and the Loneliness baseline (L-baseline), defined as the date when the loneliness responses were given (see below). A key inclusion criterion was the presence of EHR data spanning at least one year after the first EHR visit date. All participants provided informed consent for EHR linkage and survey collection. The study protocol was approved by the Institutional Review Board overseeing AoU. Our analysis was conducted under an AoU data use agreement on the secure Researcher Workbench, with all data de-identified, and results were presented in aggregate.

### Measures

#### HL

Using the OMOP CDM standardized SNOMED CT vocabulary within the AoU Researcher Workbench, we started from the ancestor concept “Hearing loss” and included all active descendants in the Condition domain to define HL diagnoses. We curated a set of 67 HL concepts (e.g., sensorineural, conductive, mixed, presbycusis; full list in Supplementary Table S1). Participants with any qualifying diagnosis recorded in their EHR were classified as HL = 1; those with no such diagnosis were classified as HL = 0 (19,20).

#### Loneliness

Loneliness was assessed by using the 8-item version of the UCLA Loneliness Scale (ULS-8) which was administered after initial enrollment, as part of the Social Determinants of Health (SDoH) survey, officially launched on November 1, 2021. This validated questionnaire (21) assesses subjective feelings of loneliness and social disconnection. The ULS-8 measure is widely used and has good psychometric properties in diverse populations (21,22). Participants rated items (e.g., “How often do you feel isolated from others?”) on a Likert scale, and we summed responses to an overall score ranging from 8 to 32, with higher scores indicating greater loneliness. We treated loneliness as a continuous variable in analyses. Because the SDoH survey allowed a ‘skip’ response option, we recoded all skipped selections as missing. We included participants who answered at least four of the eight items and computed a ULS-8 mean score across non-missing items, yielding a summary loneliness score ranging from 1 to 4 (higher scores indicating greater loneliness). This mean score was used as the continuous loneliness measure in all loneliness-related analyses. The eight item are listed in Supplementary Table S2.

#### Cognitive outcomes—incident dementia and IC

Using the OMOP CDM SNOMED CT vocabulary in the AoU Workbench, we obtained two EHR-derived outcomes. Dementia was identified by expanding from the SNOMED ancestor “Dementia” to include all active Condition-domain descendants (full code list and ICD crosswalk in Supplementary Table S3). We also examined a broader outcome that AoU terms “impaired cognition,” intended to capture milder cognitive diagnoses or prominent cognitive symptoms short of dementia. We operationalized this using a curated set of 24 OMOP standard SNOMED concepts in the Condition domain (full list in Supplementary Table S4). For both outcomes, incident status required the first qualifying diagnosis after the relevant baseline (HL-baseline for hearing-loss analyses; L-baseline for loneliness/mediation), with the event date taken as the date of first occurrence. Participants with any qualifying dementia (or impaired-cognition, respectively) code before the relevant baseline were excluded. Follow-up ran from baseline to the earliest of first qualifying diagnosis, death, or last EHR record; those without the outcome were censored at the date of death or last EHR encounter.

#### Covariates

We included a range of covariates in our models to adjust for potential confounding and to examine subgroup effects. Age (in years) was included as a continuous variable, given its strong association with both HL and dementia incidence. For analyses with HL as the exposure, age at the first EHR visit was used; for analyses with loneliness as the exposure—and for mediation analyses—age at the SDoH assessment was used. We also tested age-squared terms and categorical age bands to verify the proportional hazards assumption, with final models using the linear term. Sex/gender was included as a categorical variable (female, male, and non-binary group), as differences in both HL prevalence and dementia risk have been reported between men and women. Race/ethnicity was self-identified and categorized into mutually exclusive groups: White, Black or African American, Asian, American Indian or Alaska Native, Native Hawaiian or other Pacific Islander, more than one race, and Middle Eastern or North African. Adjusting for race/ethnicity allowed us to account for potential disparities in HL, loneliness, and dementia incidence, as well as to control for population stratification in a broad sense.

Socio-economic factors included years of education completed (treated as a continuous variable) and annual household income (self-reported in ordered categories, from <$25,000 to >$200,000). Education serves as a proxy for cognitive reserve and socio-economic status and is strongly associated with dementia risk, while income may confound the associations of interest through its influence on healthcare access, living conditions, and psychosocial stressors. We also adjusted for health behaviors, including smoking status (never, former or current, and unknown) and alcohol use (never, former or current, and unknown) at baseline.

We considered adjusting for certain comorbidities (e.g., hypertension, diabetes, depression) given their influence on dementia risk; however, many chronic conditions could lie on the causal pathway between HL and dementia (for instance, HL might lead to depression, which affects cognition). To avoid over-adjustment, we did not include comorbidities in the main models.

### Data Analysis

All analyses were conducted using R (version 4.4.0) on the AoU secure Researcher Workbench (19). We first examined participant characteristics stratified by outcome status (dementia vs. no dementia; IC vs. no impairment) and summarized continuous variables as mean (SD) and categorical variables as n (%). Between-group differences were assessed using Wilcoxon rank-sum tests for continuous variables and Pearson’s chi-squared tests for categorical variables. To minimize case loss without ad hoc imputation, we labeled missing categorical covariates as ‘Unknown’ for tabulation but excluded ‘Unknown’ observations from the primary models, and analyzed continuous covariates using available-case data. Internal consistency of the ULS-8 was assessed using Cronbach’s alpha based on item-level responses among participants with ≥4 completed items

We fit a multiple linear regression model with loneliness as the outcome and HL status as the key predictor, adjusting for all covariates (age, sex, race, education, income, smoking, alcohol). The linear model yields the adjusted mean difference in ULS-8 loneliness for participants with versus without HL (β_HL), establishing the exposure–mediator association needed for mediation analysis.

We used Cox proportional hazards models for time to incident dementia and IC. Because HL and loneliness were ascertained at different baselines—HL from the first EHR visit and loneliness from the SDoH survey—we fit separate models for each exposure (HL→dementia, HL→IC, loneliness→dementia, loneliness→IC) to respect exposure timing, avoid mixing risk sets, and maximize sample size. As sensitivity analyses, we fit time-updated Cox models treating HL as a time-varying covariate (Supplementary Methods S1); loneliness remained baseline-only due to the single assessment (23). Each Cox model was adjusted for all covariates. Participants with a record of dementia or IC at or before the respective baseline times were excluded from the corresponding models. We report hazard ratios (HRs) and 95% confidence intervals (CIs) for the associations of HL (yes vs no) and of loneliness (per unit; ULS-8 mean) with the outcomes. Proportional hazards were checked with Schoenfeld residuals; key predictors met assumptions. To determine if the associations differed by demographic or socio-economic factors, we tested interaction terms of HL and loneliness with racial groups, income, and education.

To assess whether loneliness mediates the HL→outcome association while allowing exposure–mediator interaction, we used four-way decomposition via CMAverse (*cmest*) package in R (24,25). HL (binary) was the exposure and loneliness (mean-centered continuous ULS-8) was the mediator. The outcomes were dementia and IC in separate analyses, and covariates matched those used in the aforementioned Cox models. We obtained hazard-ratio-scale estimates for the total effect, pure natural direct effect (PNDE), pure natural indirect effect (PNIE), mediated interaction, and reference interaction, and summarized mediation by the proportion mediated (pm).

All statistical tests were two-sided, with *p* < 0.05 considered statistically significant. Given the large sample size, many covariate effects reached nominal significance. We focus on those associations of substantive interest. No formal multiple comparison correction was applied for the primary hypotheses, but results were interpreted with caution and an eye to consistency with prior research. Reporting followed STROBE guidelines (26). We summarize the key study design and modeling framework in Figure 1, including the HL Cox, the Loneliness Cox, and the four-way decomposition models.

**Figure 1.**
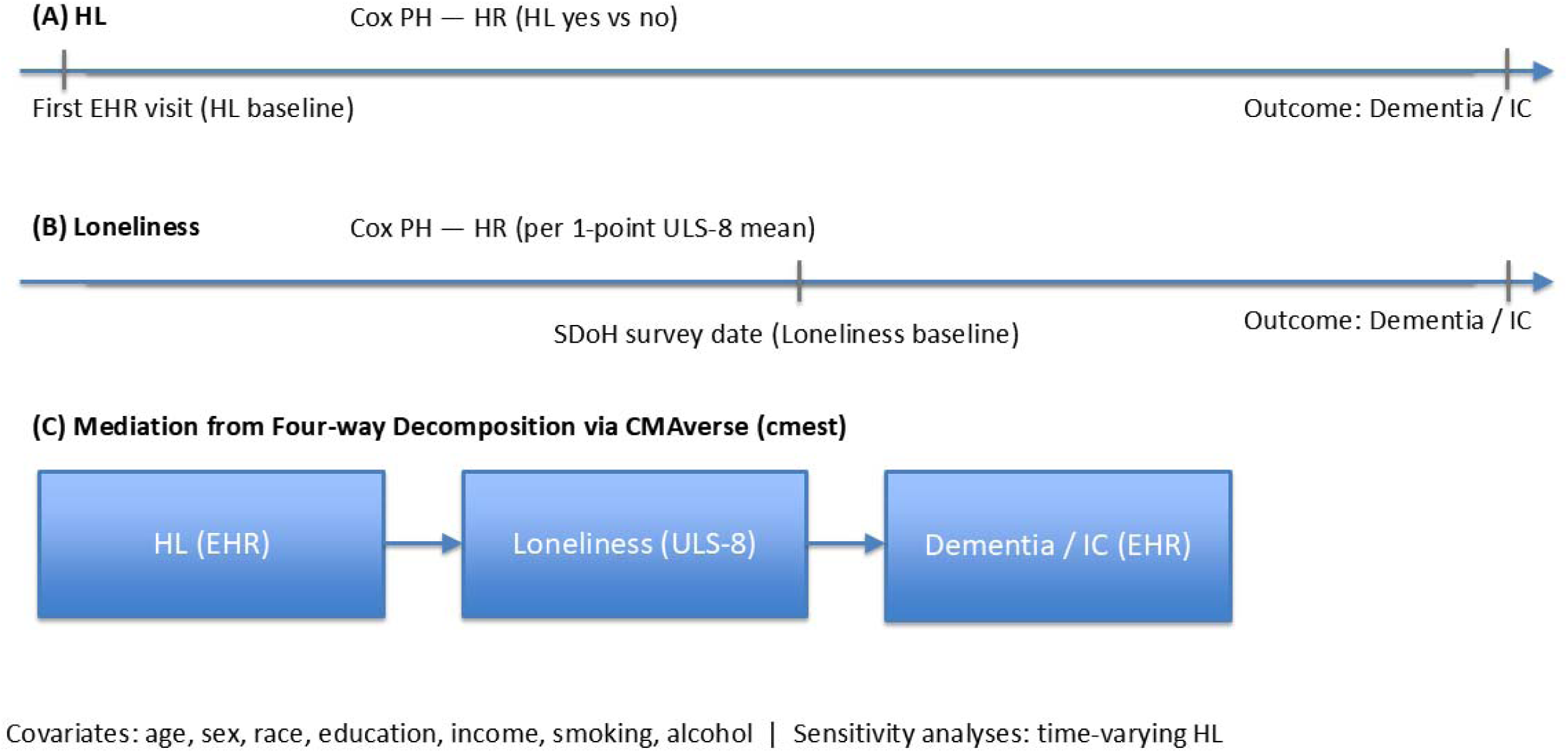
Analytical workflow and model timelines for hearing loss, loneliness, and cognitive outcomes in the All of Us cohort.

## RESULTS

### Participant Characteristics

The final analytic sample included 317,020 participants (mean overall follow-up, 11.8 years), calculated per participant from first visit date to last recorded encounter. Analyses evaluating loneliness as an exposure were restricted to those with a baseline SDoH loneliness measure. Finally, 50,807 (16.0%) had EHR-documented hearing loss, and 132,424 (41.8%) completed the SDoH survey; of these, 130,257 (98.4%) answered ≥4 items and were scored on the ULS-8. The ULS-8 demonstrated good internal consistency (Cronbach’s α = 0.87). Over follow-up, we identified 6,042 (1.9%) participants with incident dementia and 21,699 (6.8%) with incident impaired cognition; 3,034 experienced both outcomes, either concurrently or sequentially. An additional 162 participants had impaired cognition prior to the first EHR visit were excluded from incident impaired-cognition analyses.

Table 1 displays the characteristics of participants who did vs. did not develop dementia during follow-up. Out of 317,020 participants, 6,042 (1.9%) were diagnosed with dementia. Dementia cases were significantly older (mean age 53.62 ± 16.06 years) compared with non-cases (44.00 ± 16.63 years; *p* < 0.001). Dementia cases were more likely to be male (51% vs 38%; *p* < 0.001) and to have hearing loss (37% vs 16%; *p* < 0.001). Mean loneliness scores were higher among dementia cases (2.06 ± 0.69) than non-cases (1.90 ±0.65; *p* < 0.001). Smoking prevalence was greater in dementia cases (52% current/former) compared with non-cases (41%; *p* < 0.001), and dementia cases more frequently reported never drinking alcohol (13% vs 10%; *p* < 0.001). Lower educational attainment was more common among dementia cases, with 35% having ≤12 years of education compared with 27.7% of non-cases (*p* < 0.001). Income levels were also lower among cases; 62% of dementia cases had an annual household income ≤$42,500 compared with 48.2% of non-cases (*p* < 0.001).

**Table 1.** The characteristics of participants who did versus did not develop dementia during follow-up.

| Characteristic | N | Dementia |  | p-value<sup>2</sup> |
| --- | --- | --- | --- | --- |
|  |  | No<br>N = 310,978<sup>1</sup> | Yes<br>N = 6,042<sup>1</sup> |  |
| **Hearing loss** | 317,020 |  |  | <0.001 |
| No |  | 262,398 (84%) | 3,815 (63%) |  |
| Yes |  | 48,580 (16%) | 2,227 (37%) |  |
| **Gender** | 312,264 |  |  | <0.001 |
| Female |  | 189,887 (62%) | 2,922 (49%) |  |
| Male |  | 115,565 (38%) | 2,994 (51%) |  |
| Non-Binary |  | 887 (0.3%) | <20 |  |
| Unknown |  | 4,639 | 117 |  |
| **Race** | 263,844 |  |  |  |
| American Indian or Alaska Native |  | 3,616 (1.4%) | 115 (2.3%) |  |
| Asian |  | 8,673 (3.4%) | 78 (1.6%) |  |
| Black or African American |  | 57,947 (22%) | 943 (19%) |  |
| Middle Eastern or North African |  | 1,732 (0.7%) | 27 (0.5%) |  |
| More than one population |  | 13,027 (5.0%) | 276 (5.6%) |  |
| Native Hawaiian or Other Pacific Islander |  | 262 (0.1%) | <20 |  |
| White |  | 173,625 (67%) | 3,521 (71%) |  |
| Unknown |  | 52,096 | 1,080 |  |
| **Education years** | 309,520 |  |  | <0.001 |
| 1 |  | 401 (0.1%) | 22 (0.4%) |  |
| 4 |  | 2,421 (0.8%) | 122 (2.1%) |  |
| 6.5 |  | 6,456 (2.1%) | 244 (4.2%) |  |
| 10 |  | 17,427 (5.7%) | 371 (6.3%) |  |
| 12 |  | 57,170 (19%) | 1,275 (22%) |  |
| 14 |  | 79,650 (26%) | 1,593 (27%) |  |
| 16 |  | 71,715 (24%) | 1,113 (19%) |  |
| 18 |  | 68,414 (23%) | 1,126 (19%) |  |
| Unknown |  | 7,324 | 176 |  |
| **Income (dollar)** | 254,206 |  |  | <0.001 |
| 5000 |  | 39,240 (16%) | 846 (19%) |  |
| 17500 |  | 34,968 (14%) | 979 (22%) |  |
| 30000 |  | 21,142 (8.5%) | 460 (10%) |  |
| 42500 |  | 24,232 (9.7%) | 498 (11%) |  |
| 62500 |  | 33,348 (13%) | 565 (13%) |  |
| 87500 |  | 26,305 (11%) | 339 (7.7%) |  |
| 125000 |  | 33,034 (13%) | 394 (8.9%) |  |
| 175000 |  | 15,639 (6.3%) | 152 (3.4%) |  |
|  |  | No<br>N = 310,978<sup>1</sup> | Yes<br>N = 6,042<sup>1</sup> |  |
| 225000 |  | 21,875 (8.8%) | 190 (4.3%) |  |
| Unknown |  | 61,195 | 1,619 |  |
| **Smoking** | 307,240 |  |  | <0.001 |
| Never |  | 178,879 (59%) | 2,825 (48%) |  |
| Former or current |  | 122,497 (41%) | 3,039 (52%) |  |
| Unknown |  | 9,602 | 178 |  |
| **Drinking** | 309,948 |  |  | <0.001 |
| Never |  | 30,335 (10.0%) | 748 (13%) |  |
| Former or current |  | 273,724 (90%) | 5,141 (87%) |  |
| Unknown |  | 6,919 | 153 |  |
| **Age (at first EHR visit)** | 317,020 | 44.00 (16.63) | 53.62 (16.06) | <0.001 |
| **Loneliness-ULS8 score** | 130,257 | 1.90 (0.65) | 2.06 (0.69) | <0.001 |
| Unknown |  | 182,233 | 4,530 |  |

<sup>1</sup>Mean (SD); n (%)<sup>2</sup>Wilcoxon rank sum test; Pearson's Chi-squared test

Table 2 presents analogous data for participants who did vs did not develop IC. A total of 21,699 participants (6.8%) were diagnosed with IC. These individuals were older (50.63 ± 15.24 years) than those without a diagnosis (43.71 ± 16.67 years; p < 0.001). HL prevalence was markedly higher among IC cases (40% vs 14%; p < 0.001), and mean loneliness scores were also greater (2.05 ± 0.69; p < 0.001). IC cases were slightly less likely to be female (61% vs 62%; p = 0.016) and more likely to be White (76% vs 66%; p < 0.001). Lower educational attainment was less frequent among cases, with 23.7% having ≤12 years of education compared with 28% of non-cases (p < 0.001). A higher proportion of IC cases reported annual household incomes ≤$42,500 (50.7% vs 48%; p < 0.001). Smoking prevalence was also higher in cases (46% current/former) than in non-cases (40%; p < 0.001), while drinking patterns showed smaller differences.

**Table 2.** The characteristics of participants who did versus did not develop impaired cognition during follow-up.

| Characteristic | N | Incident IC (pre-IC excluded) |  | p-value<sup>2</sup> |
| --- | --- | --- | --- | --- |
|  |  | No<br>N = 295,159<sup>1</sup> | Yes<br>N = 21,699<sup>1</sup> |  |
| **Hearing loss** | 316,858 |  |  | <0.001 |
| No |  | 253,028 (86%) | 13,061 (60%) |  |
| Yes |  | 42,131 (14%) | 8,638 (40%) |  |
| **Gender** | 312,104 |  |  | 0.016 |
| Female |  | 179,681 (62%) | 13,029 (61%) |  |
| Male |  | 110,224 (38%) | 8,274 (39%) |  |
| Non-Binary |  | 847 (0.3%) | 49 (0.2%) |  |
| Unknown |  | 4,407 | 347 |  |
| **Race** | 263,701 |  |  | <0.001 |
| American Indian or Alaska Native |  | 3,533 (1.4%) | 192 (1.0%) |  |
| Asian |  | 8,426 (3.4%) | 324 (1.8%) |  |
| Black or African American |  | 56,115 (23%) | 2,767 (15%) |  |
| Middle Eastern or North African |  | 1,634 (0.7%) | 124 (0.7%) |  |
| More than one population |  | 12,308 (5.0%) | 983 (5.3%) |  |
| Native Hawaiian or Other Pacific Islander |  | 247 (0.1%) | <20 |  |
| White |  | 162,965 (66%) | 14,066 (76%) |  |
| Unknown |  | 49,931 | 3,226 |  |
| **Education years** | 309,362 |  |  | <0.001 |
| 1 |  | 382 (0.1%) | 41 (0.2%) |  |
| 4 |  | 2,326 (0.8%) | 215 (1.0%) |  |
| 6.5 |  | 6,198 (2.2%) | 501 (2.4%) |  |
| 10 |  | 16,917 (5.9%) | 870 (4.1%) |  |
| 12 |  | 54,918 (19%) | 3,496 (16%) |  |
| 14 |  | 75,049 (26%) | 6,140 (29%) |  |
| 16 |  | 67,857 (24%) | 4,944 (23%) |  |
| 18 |  | 64,439 (22%) | 5,069 (24%) |  |
| Unknown |  | 7,073 | 423 |  |
| **Income (dollar)** | 254,083 |  |  | <0.001 |
| 5000 |  | 38,011 (16%) | 2,051 (12%) |  |
| 17500 |  | 32,812 (14%) | 3,115 (18%) |  |
| 30000 |  | 19,912 (8.4%) | 1,670 (9.7%) |  |
| 42500 |  | 22,809 (9.6%) | 1,906 (11%) |  |
| 62500 |  | 31,275 (13%) | 2,622 (15%) |  |
| 87500 |  | 24,769 (10%) | 1,864 (11%) |  |
| 125000 |  | 31,301 (13%) | 2,117 (12%) |  |
| 175000 |  | 14,957 (6.3%) | 831 (4.8%) |  |
|  |  | No<br>N = 295,159<sup>1</sup> | Yes<br>N = 21,699<sup>1</sup> |  |
| 225000 |  | 20,959 (8.9%) | 1,102 (6.4%) |  |
| Unknown |  | 58,354 | 4,421 |  |
| **Smoking** | 307,080 |  |  | <0.001 |
| Never |  | 170,175 (60%) | 11,443 (54%) |  |
| Former or current |  | 115,813 (40%) | 9,649 (46%) |  |
| Unknown |  | 9,171 | 607 |  |
| **Drinking** | 309,787 |  |  | 0.003 |
| Never |  | 29,054 (10%) | 2,007 (9.4%) |  |
| Former or current |  | 259,447 (90%) | 19,279 (91%) |  |
| Unknown |  | 6,658 | 413 |  |
| **Age (at first EHR visit)** | 316,858 | 43.71 (16.67) | 50.63 (15.24) | <0.001 |
| **Loneliness-ULS8 score** | 130,205 | 1.90 (0.65) | 2.05 (0.69) | <0.001 |
| Unknown |  | 174,344 | 12,309 |  |

<sup>1</sup>Mean (SD); n (%)<sup>2</sup>Wilcoxon rank sum test; Pearson's Chi-squared test

### The Association Between HL and Loneliness

We examined the cross-sectional relationship between HL and loneliness using a multiple linear regression model, adjusting for all covariates (Supplementary Table S5). Participants with HL reported a 0.28-point higher ULS-8 total score (β on the ULS-8 mean = 0.035, SE = 0.005, p < 0.001). As an effect-size benchmark, this hearing-loss coefficient is roughly equivalent in magnitude to ∼5.3 years of age difference (age β = −0.0067 per year on the mean; ≈ −0.053 per year on the total), albeit in the opposite direction. All coefficients are unstandardized. Compared with White participants, Black/African American participants reported lower loneliness (β = −0.050 on the mean; ≈ −0.40 on the total; p < 0.001), and multiracial participants reported higher scores (β = 0.077; ≈ +0.62 total; p < 0.001). American Indian/Alaska Native participants showed a lower mean score (β = −0.048; ≈ −0.38 total) but this was borderline (p = 0.063). Non-binary gender was strongly associated with higher loneliness (β = 0.319; ≈ +2.55 total; p < 0.001), and males reported slightly higher scores (β = 0.011; ≈ +0.09 total; p = 0.006). Higher education was positively associated with loneliness (β = 0.0033 per year; ≈ +0.026 total; p = 0.002), while income was inversely associated (β = −2.427×10^⁻^□ per dollar; ≈ −0.19 on the total per $10,000; p < 0.001). Smoking was positively associated (β = 0.053; ≈ +0.42 total; p < 0.001), whereas alcohol use was not significant (β = 0.003; ≈ +0.03 total; p = 0.768). In summary, HL was independently associated with greater loneliness, fulfilling a key requirement for mediation (i.e., the exposure is associated with the mediator), and supporting the hypothesis that loneliness may mediate the pathway from HL to cognitive outcomes.

### HL and Loneliness as Predictors of Dementia

In our Cox proportional hazards models, we observed that HL significantly increased the risk of developing dementia. The adjusted Cox model revealed a HR of 1.29 (p < 0.001), indicating a notably higher risk of dementia in individuals with HL compared to those without it. Participants with HL had a 29% higher hazard of developing dementia compared with those without HL. Similarly, loneliness was associated with increased dementia hazard. Per 1-point increase in ULS-8 mean (item-average), HR = 1.41 (95% CI 1.12–1.75, p = 0.003), or HR ≈ 1.04 per 1-point on the ULS-8 total. As an age benchmark, HL corresponds to roughly +4.4 years of age in dementia hazard (age HR≈1.06/year). A 1-point increase in ULS-8 mean corresponds to about +5.9 years (95% CI +1.9–9.6), and each 1-point increase in the ULS-8 total is roughly +0.67 years. Alcohol use was associated with lower hazard (HR in the loneliness model = 0.37, 95% CI 0.21–0.67, p = 0.001). Table 3 presents the complete Cox regression results for the relationship between HL and dementia, as well as loneliness and dementia.

**Table 3.** (a). Cox proportional hazards regression results of hearing loss on dementia, adjusted by all covariates, with baseline at first EHR-recorded visit date (N = 208,665; events = 3,502); (b). Cox proportional hazards regression results of loneliness (ULS-8 mean, item-average) on dementia, adjusted for all covariates, baseline at loneliness survey date (N = 100,603; events = 191)

| Variable | HR | SE | 95% CI | p-value |
| --- | --- | --- | --- | --- |
| **Model 1: Hearing loss on dementia** |  |  |  |  |
| Hearing loss (yes vs no) | 1.294 | 0.037 | 1.204, 1.390 | <0.001 |
| Age (per year) | 1.059 | 0.001 | 1.056, 1.062 | <0.001 |
| Gender |  |  |  |  |
| Male vs Female | 1.494 | 0.035 | 1.396, 1.600 | <0.001 |
| Non-binary vs Female | 2.489 | 0.410 | 1.115, 5.557 | 0.026 |
| Race/Ethnicity (ref: White) |  |  |  |  |
| American Indian/Alaska Native | 2.084 | 0.129 | 1.617, 2.685 | <0.001 |
| Asian | 1.082 | 0.143 | 0.818, 1.432 | 0.579 |
| Black/African American | 0.944 | 0.050 | 0.855, 1.041 | 0.247 |
| Middle Eastern/North African | 0.873 | 0.290 | 0.494, 1.541 | 0.639 |
| More than one population | 1.463 | 0.076 | 1.261, 1.697 | <0.001 |
| Native Hawaiian/Other Pacific Islander | 0.000005 | 322.9 | 7.33e-281, 3.39e+269 | 0.970 |
| Education (per year) | 0.964 | 0.008 | 0.949, 0.979 | <0.001 |
| Income (per dollar) | 1.000 | 0.000 | 1.000, 1.000 | <0.001 |
| Smoking (ever/current vs never) | 1.154 | 0.035 | 1.076, 1.237 | <0.001 |
| Alcohol use (ever/current vs never) | 0.710 | 0.068 | 0.622, 0.812 | <0.001 |
| **Model 2: Loneliness on dementia** |  |  |  |  |
| Loneliness score (ULS-8 mean) | 1.405 | 0.114 | 1.124, 1.755 | 0.003 |
| Age (per year) | 1.064 | 0.007 | 1.049, 1.078 | <0.001 |
| Gender |  |  |  |  |
| Male vs Female | 1.305 | 0.149 | 0.975, 1.747 | 0.073 |
| Non-binary vs Female | 0.000000913 | 2114.0 | 0.000, Inf | 0.995 |
| Race/Ethnicity (ref: White) |  |  |  |  |
| American Indian/Alaska Native | 0.000000308 | 1923.0 | 0.000, Inf | 0.994 |
| Asian | 1.524 | 0.514 | 0.557, 4.171 | 0.412 |
| Black/African American | 1.178 | 0.268 | 0.696, 1.992 | 0.542 |
| Middle Eastern/North African | 1.391 | 1.006 | 0.194, 9.984 | 0.743 |
| More than one population | 1.431 | 0.365 | 0.700, 2.928 | 0.326 |
| Native Hawaiian/Other Pacific Islander | 0.000000265 | 6982.0 | 0.000, Inf | 0.998 |
| Education (per year) | 0.982 | 0.038 | 0.911, 1.058 | 0.628 |
| Income (per dollar) | 1.000 | 0.00000148 | 1.000, 1.000 | 0.003 |
| Smoking (ever/current vs never) | 1.190 | 0.152 | 0.883, 1.604 | 0.254 |
| Alcohol use (ever/current vs never) | 0.375 | 0.293 | 0.211, 0.666 | 0.001 |

In interaction analyses, the association between HL and dementia was significantly modified by race and income, but not by education (Supplementary Table S6). Compared with White participants, this association was stronger among Black participants (HR for interaction = 1.76, 95% CI = 1.45–2.13, p < 0.001), while no significant interactions were observed for other racial groups. The interaction between HL and income was positive and statistically significant (HR ≈ 1.000 per unit income, p = 0.030), indicating a slightly stronger association at higher income levels. No evidence of effect modification by education was found (p = 0.245). Although loneliness was significantly associated with an increased risk of dementia, there was no evidence of effect modification by race or education (all interaction p > 0.14), but a very small income-by-loneliness interaction was detected (HR ≈ 1.000, p = 0.030), indicating a slightly weaker association at higher income (≈1.7% attenuation per $10,000; Supplementary Table S7).

### HL and Loneliness as Predictors of IC

We observed similar associations for IC. HL significantly increased the hazard of developing IC (HR=1.65; p < 0.001). Similarly, loneliness was a significant predictor of IC. Per 1-point increase in the ULS-8 mean (item-average), the HR was 1.49 (95% CI 1.36–1.62, p < 0.001), ≈ 1.05 per 1-point on the ULS-8 total. As an age benchmark, the HL effect corresponds to roughly +12.5 years of age (95% CI +11.6–13.4), and a 1-point increase in ULS-8 mean corresponds to about +14.3 years (95% CI +11.1–17.5). Table 4 presents the Cox regression results for the relationship between HL and IC, as well as loneliness and IC. Although HL was strongly associated with increased risk of IC, the magnitude of this association varied by race: compared with White participants, it was larger among Asian (interaction HR = 1.42, p = 0.013) and Black/African American participants (interaction HR = 1.52, p < 0.001). By contrast, interactions with income and education were not significant (p = 0.59 and p = 0.24, respectively; Supplementary Table S8). Although loneliness (per 1-point increase in ULS-8 mean) was strongly associated with increased risk of IC, this association did not vary by race (all interaction p ≥ 0.14). By contrast, we detected a potential interaction with income (HR≈1, p = 0.033; the loneliness–IC association was slightly weaker among participants with higher income), whereas the interaction with education was not significant (p = 0.36; Supplementary Table S9).

**Table 4.** (a). Cox proportional hazards regression of hearing loss on impaired cognition, adjusted by all covariates, baseline at first visit date (N = 208,315; events = 14,168); (b). Cox proportional hazards regression of loneliness (ULS-8 mean, item-average) on impaired cognition, adjusted by all covariates, baseline at loneliness survey date (N = 81,606; events = 1,180)

| Variable | HR | SE | 95% CI | p-value |
| --- | --- | --- | --- | --- |
| **Model 1: Hearing loss on impaired cognition** |  |  |  |  |
| Hearing loss (yes vs no) | 1.653 | 0.01791 | 1.596, 1.712 | <0.001 |
| Age (per year) | 1.041 | 0.00064 | 1.040, 1.043 | <0.001 |
| Gender |  |  |  |  |
| Male vs Female | 0.867 | 0.01766 | 0.837, 0.897 | <0.001 |
| Non-binary vs Female | 2.299 | 0.16310 | 1.670, 3.165 | <0.001 |
| Race/Ethnicity (ref: White) |  |  |  |  |
| Asian | 0.904 | 0.11360 | 0.724, 1.130 | 0.377 |
| Black/African American | 1.041 | 0.01013 | 1.021, 1.062 | <0.001 |
| Middle Eastern/North African | 1.158 | 0.14870 | 0.865, 1.550 | 0.324 |
| More than one population | 0.976 | 0.03883 | 0.905, 1.053 | 0.535 |
| Native Hawaiian/Other Pacific Islander | 0.657 | 0.18110 | 0.460, 0.936 | 0.020 |
| Education (per year) | 1.000 | 0.04167 | 0.922, 1.085 | 0.915 |
| Income (per dollar) | 1.000 | 1.67×10<sup>-4</sup> | 1.000, 1.000 | <0.001 |
| Smoking (ever/current vs never) | 0.998 | 0.01752 | 0.965, 1.033 | 0.925 |
| Alcohol use (ever/current vs never) | 0.936 | 0.03898 | 0.867, 1.010 | 0.087 |
| **Model 2: Loneliness on impaired cognition** |  |  |  |  |
| Loneliness score (ULS-8 mean) | 1.485 | 0.045 | 1.358, 1.623 | <0.001 |
| Age at loneliness (per year) | 1.028 | 0.002 | 1.023, 1.033 | <0.001 |
| Gender |  |  |  |  |
| Male vs Female | 1.032 | 0.061 | 0.915, 1.163 | 0.609 |
| Non-binary vs Female | 1.245 | 0.504 | 0.464, 3.345 | 0.663 |
| Race/Ethnicity (ref: White) |  |  |  |  |
| American Indian/Alaska Native | 1.687 | 0.336 | 0.872, 3.261 | 0.120 |
| Asian | 0.537 | 0.305 | 0.295, 0.976 | 0.041 |
| Black/African American | 0.769 | 0.123 | 0.604, 0.978 | 0.032 |
| Middle Eastern/North African | 1.660 | 0.356 | 0.826, 3.336 | 0.155 |
| More than one population | 1.133 | 0.151 | 0.844, 1.522 | 0.407 |
| Native Hawaiian/Other Pacific Islander | 0.00001 | 380.7 | 0.000, Inf | 0.977 |
| Education (per year) | 0.975 | 0.016 | 0.946, 1.006 | 0.112 |
| Income (per dollar) | 1.000 | 5.44×10<sup>-4</sup> | 1.000, 1.000 | 0.001 |
| Smoking (ever/current vs never) | 1.090 | 0.061 | 0.967, 1.229 | 0.159 |
| Alcohol use (ever/current vs never) | 0.817 | 0.163 | 0.593, 1.125 | 0.215 |

Consistent with these estimates, Kaplan–Meier curves show shorter time to dementia and IC among those with HL or greater loneliness (Figs. 2–3), and forest plots provide the corresponding adjusted HRs with 95% CIs (Figs. 4–5).

**Figure 2.**
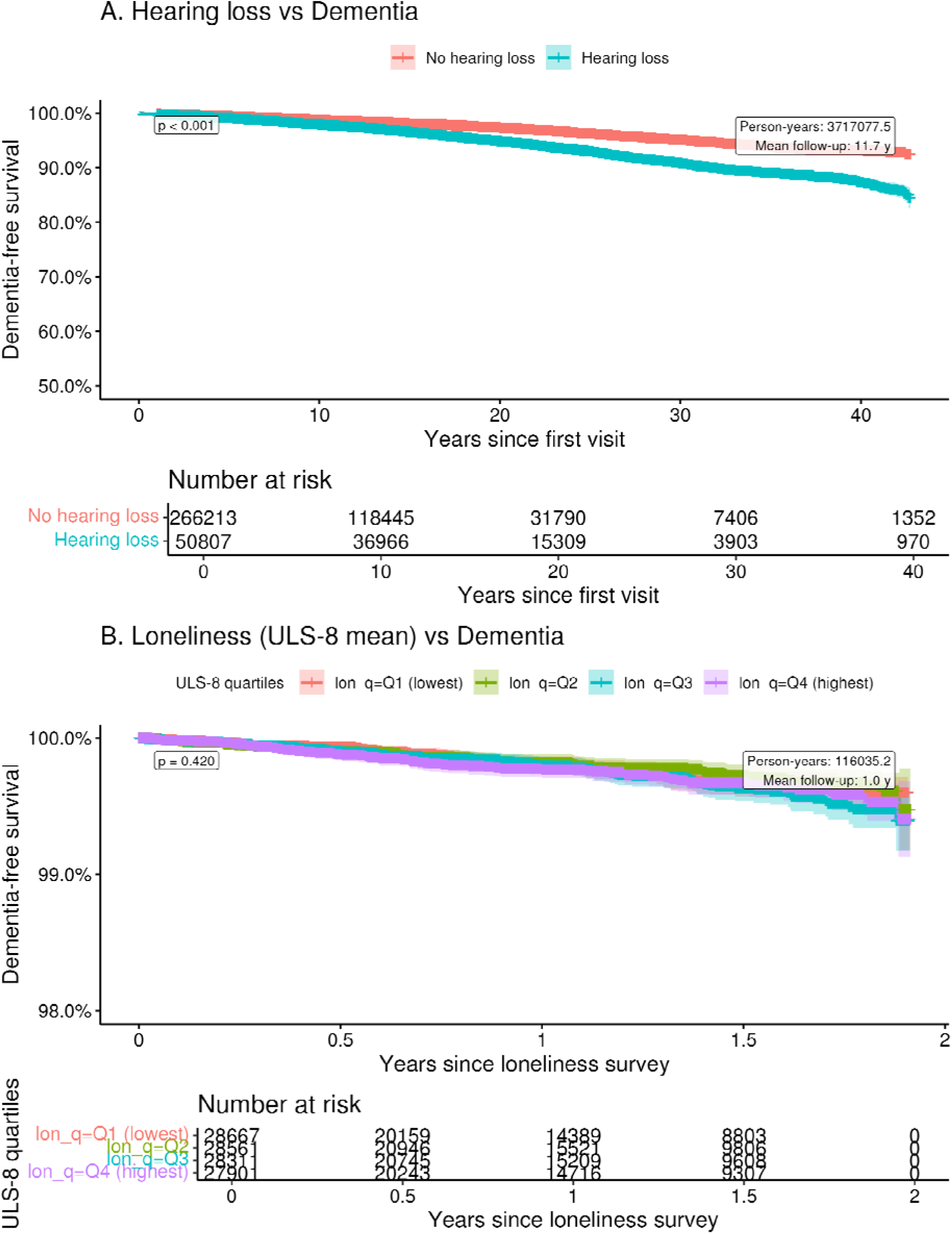
Kaplan–Meier curves for dementia-free survival by baseline hearing loss status. Shaded areas indicate 95% confidence intervals; p-value from log-rank test. Numbers at risk are shown below the plot.

**Figure 3.**
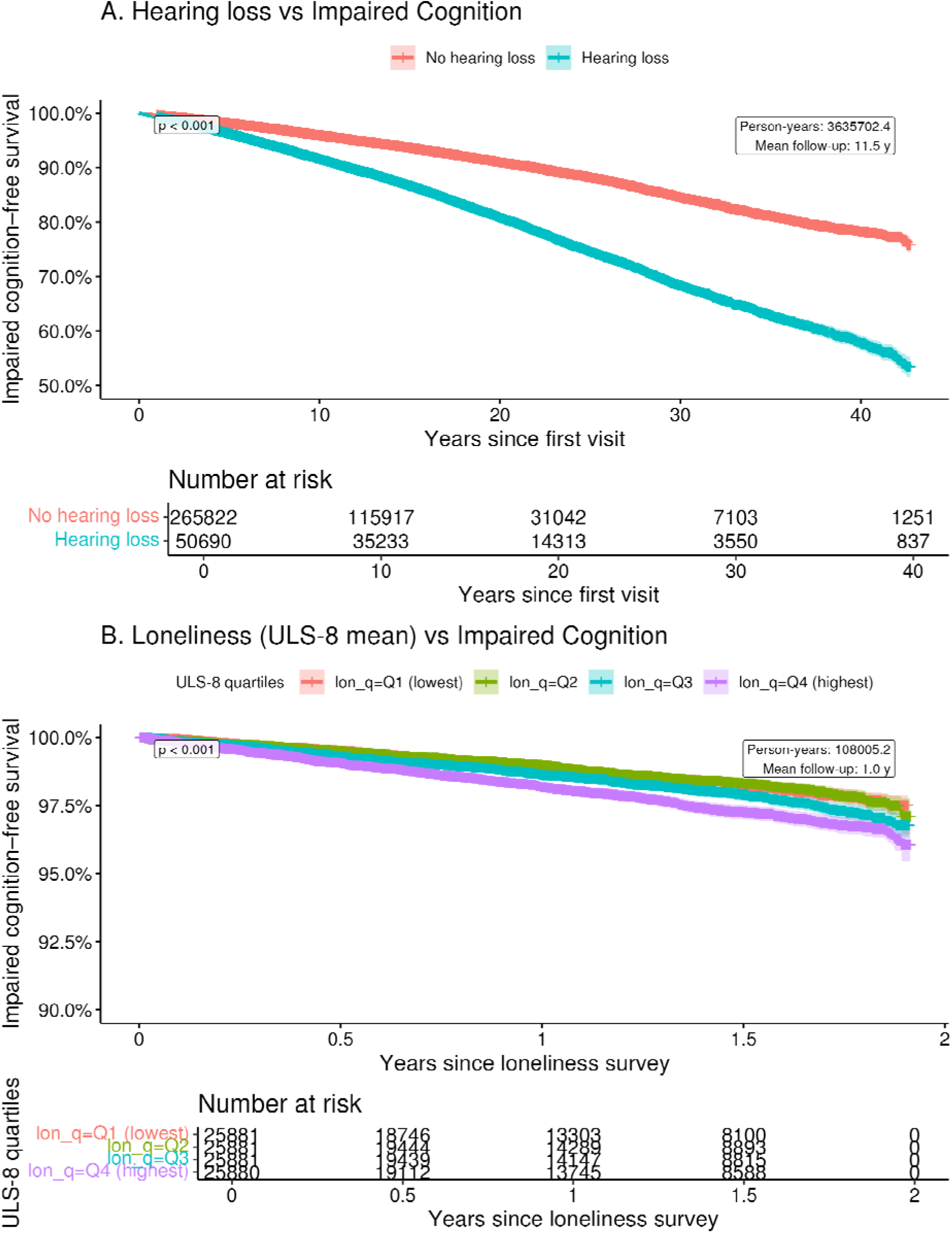
Kaplan–Meier curves for impaired cognition-free survival by baseline hearing loss status. Shaded areas indicate 95% confidence intervals; p-value from log-rank test. Numbers at risk are shown below the plot.

**Figure 4.**
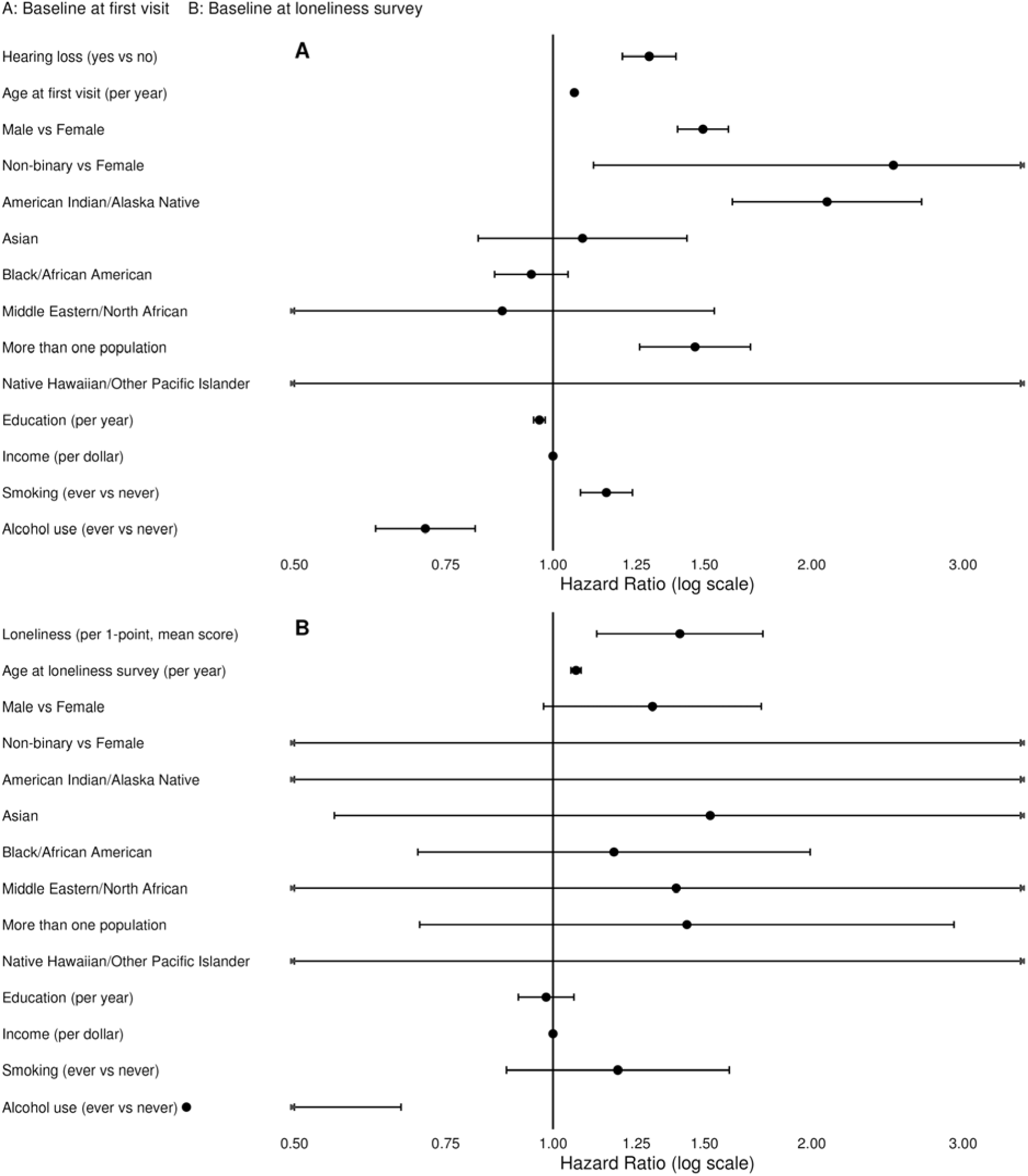
Forest plots showing hazard ratios (HR) for all variables included in multivariable models with dementia as outcome (A. HL; B. Loneliness)

**Figure 5.**
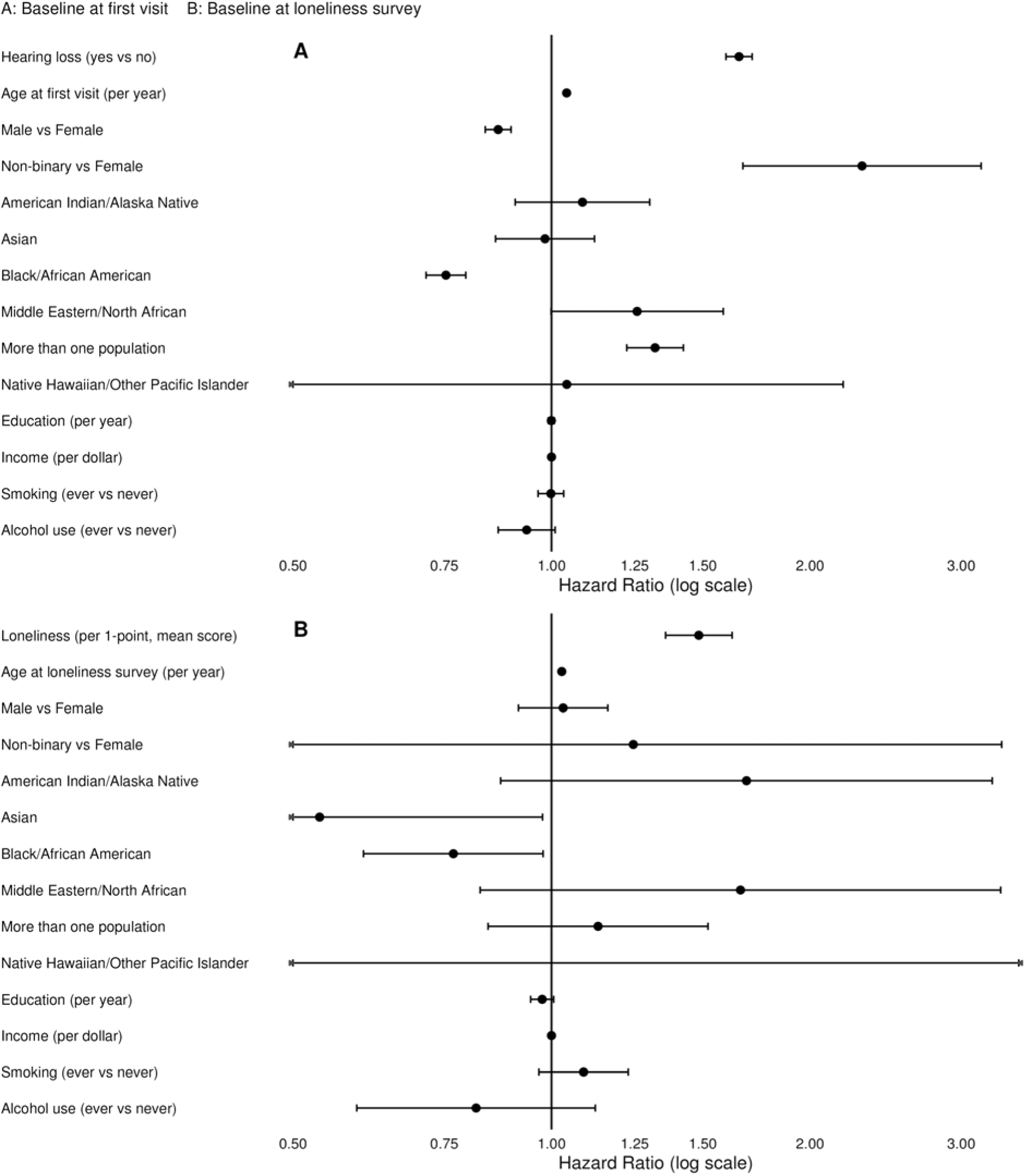
Forest plots showing hazard ratios (HR) for all variables included in multivariable models with impaired cognition as outcome (A. HL; B. Loneliness)

### Sensitivity analysis: Time-Varying Cox Analysis of HL on cognitive outcomes

As a sensitivity analysis, we re-fit Cox models with HL coded as time-varying. Estimates were similar to the baseline-exposure models (Supplementary Materials Results SR1 and Tables S10–S11).

### Mediation Analysis: Loneliness as a Mediator of HL Effect

Full four-way outputs (including interaction components) are provided in Table 5. Because we did not pre-specify hypotheses about interaction between HL and loneliness, we restrict main-text reporting to mediation components.

**Table 5.**
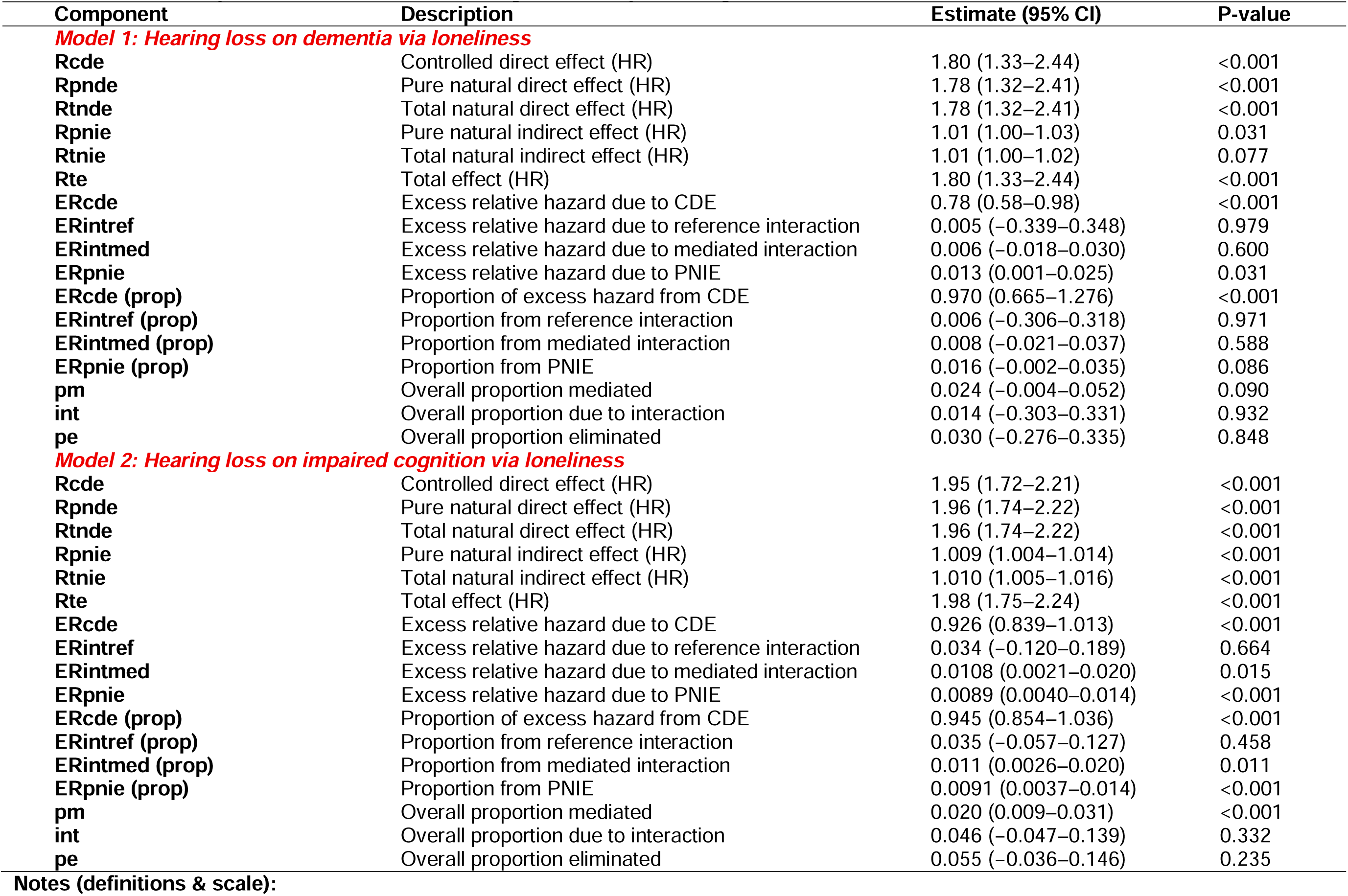
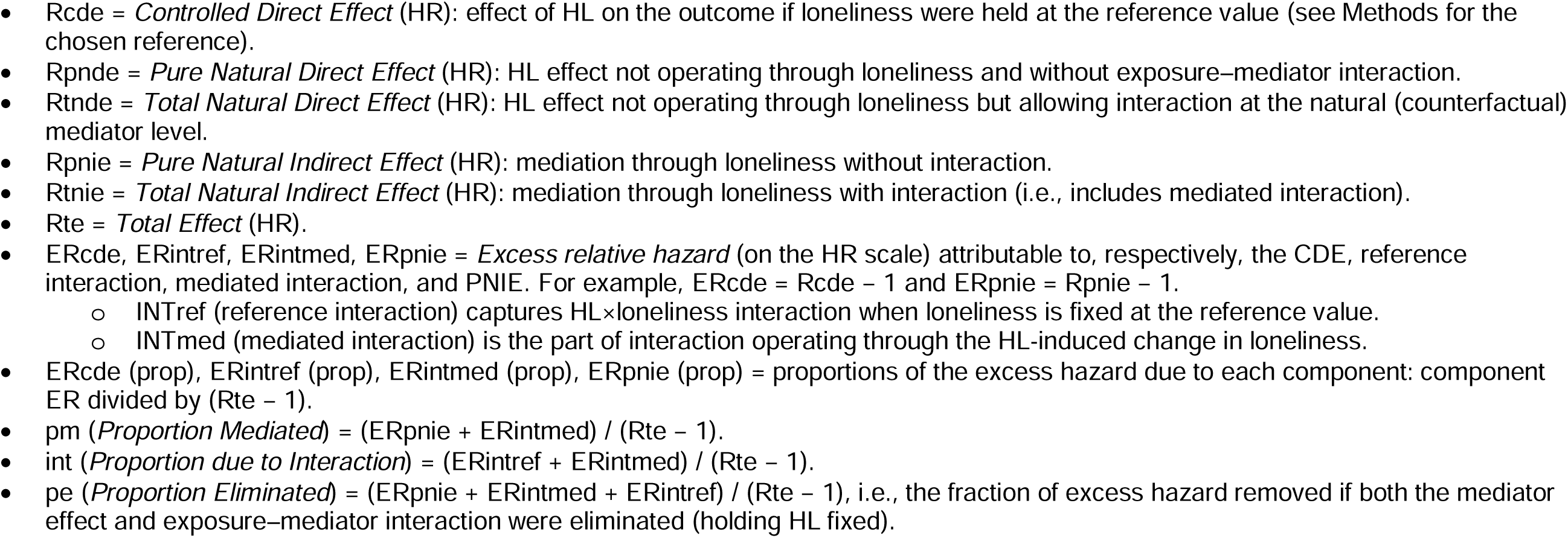
(a). Four-way decomposition of the effect of hearing loss on dementia via loneliness (N = 85,979; events = 185); (b). Four-way decomposition of the effect of hearing loss on impaired cognition via loneliness (N = 81,205; events = 1,162)

The total effect of HL on incident dementia was significant (Rte = 1.80, p < 0.001) and was driven almost entirely by the natural direct effect (Rcde = 1.80, p < 0.001; ERcde(prop) ≈ 0.97). The indirect pathway via loneliness was not significant (Rtnie = 1.011, p = 0.077), yielding an overall proportion mediated of 2.4% (pm = 0.024, p = 0.090). For incident IC, the total effect was likewise significant (Rte = 1.98, p < 0.001) and largely direct (Rcde = 1.95, p < 0.001), while the indirect effect through loneliness was small but statistically significant (Rtnie = 1.01, p = 0.0003), yielding an overall proportion mediated of 2.0% (pm = 0.020, p = 0.0004).

## DISCUSSION

In this large, demographically diverse U.S. cohort (mean age = 44.18 years, SD = 16.67), both HL and loneliness were associated with increased risk of IC and dementia. These findings extend prior evidence that midlife HL is a modifiable dementia risk factor (1) and that loneliness harms late-life brain health (11), and they show that these relationships hold in a contemporary, broadly representative U.S. population. Consistent with the slightly larger effects for IC than for dementia, the results also suggest that psychosocial factors such as loneliness may shape cognitive aging at earlier stages. Mediation analyses indicated that loneliness explained a small but non-zero fraction of the HL–IC association.

The adjusted HRs we observed for HL (on the order of 1.29–1.65, based on external benchmarks and our models) are in line with meta-analytic estimates in the literature (1). There are several implications of our findings. HL is common in later life—about 16% of our cohort had a diagnosis of HL in their EHR record, and prevalence rises with age—and it is potentially modifiable through rehabilitative interventions, notably hearing aids and, for severe-to-profound loss, cochlear implants. Recent evidence links these devices with slower cognitive decline and reduced dementia risk (27,28). To the extent that the association between HL and dementia is causal, addressing HL could delay or prevent a portion of dementia cases. Notably, a U.S. randomized controlled trial—the ACHIEVE trial—examined the effect of a hearing intervention on cognitive decline. While the primary analysis in the general sample revealed no differences, a pre-specified subgroup of participants at higher dementia risk showed a 48% slower rate of cognitive decline over 3 years with a comprehensive hearing intervention (including hearing aid fitting and support) compared to a control group (29). This suggests that treating HL can indeed benefit cognitive outcomes, especially in those who are vulnerable. Our observational findings complement this by highlighting hearing impairment as a strong risk marker. Together, they strengthen the rationale for early hearing assessment and rehabilitation as part of dementia prevention strategies.

Mechanistically, our results lend support to the “cascade hypothesis” wherein HL leads to cognitive decline through multiple pathways. We explicitly explored one such pathway—loneliness—and found that it is operative but explains only a small fraction of the HL effect. Using four-way decomposition, we found the proportion mediated by loneliness was ∼2.4% for dementia (pm = 0.024, p = 0.090), i.e., not statistically significant, and ∼2% for IC (pm = 0.020, p = 0.0004), a small but statistically significant share. While not the dominant pathway, loneliness does play a mediating role: HL may lead to feelings of loneliness, which then modestly elevate cognitive decline risk. Conceptually, this supports the idea that social engagement is important for cognitive resilience. However, loneliness mediated only ∼2% of the HL effect on IC, consistent with a recent systematic review reporting limited evidence that social isolation mediates the HL–cognition association (18). In fact, only one longitudinal study in that review explicitly tested mediation and found no significant effect of isolation. Our quantitatively small mediation effect estimate implies that interventions solely aimed at reducing loneliness among those with HL might have a relatively minor impact on dementia incidence—the bulk of the risk must be addressed by restoring hearing (sensory input) or other means. That said, even a small mediated effect can be meaningful at a population level, given the large number of people with HL. Overall, this pattern supports a combined strategy of hearing rehabilitation and social-engagement support.

We confirmed that loneliness independently predicts risk of dementia and IC, consistent with growing evidence that loneliness/social isolation harm brain health—for example, a recent meta-analysis reported ≈30% higher dementia risk among people with loneliness (11). In adjusted hazard models, loneliness remained a significant predictor, potentially reflecting pathways noted previously—chronic stress, vascular dysregulation, and reduced cognitive engagement—though we did not measure these mechanisms (12). Importantly, loneliness may also reflect prodromal dementia—preclinical withdrawal or isolation—complicating causal interpretation (16). Although we excluded baseline cognitive-impairment diagnoses, prodromal effects cannot be fully excluded.

We pre-specified a working causal model in which HL increases loneliness, which in turn contributes to dementia risk. Although direct biological pathways from loneliness to auditory pathology are uncertain, recent prospective evidence indicates that loneliness predicts incident HL, suggesting potential bi-directionality (30). In our data, participants with HL reported higher loneliness at baseline, consistent with the hypothesized hearing→loneliness pathway; moreover, hearing status was ascertained from longitudinal EHRs, which often predate the loneliness survey, lending partial temporal support. That said, common causes—such as personality traits or depressive symptoms—could predispose individuals both to feel lonely and to have worse or unaddressed hearing, and residual confounding may remain despite adjustment for demographic and behavioral covariates. Future work with longer post-survey follow-up, repeated psychosocial assessments, and designs that better separate timing (e.g., time-updated models or quasi-experimental approaches) will be important to clarify directionality and mechanisms.

### Subgroup Analyses and Generalizability

Leveraging the representativeness of the AoU, we tested effect modification by race/ethnicity, education, and income in Cox models for HL and for loneliness. For dementia, the HL association was stronger among Black participants and modestly stronger at higher income, with no interaction by education; and in terms of loneliness, we found no interactions by race or education, but a very small income-by-loneliness interaction consistent with a slightly weaker association at higher income. For IC, the HL association varied by race—stronger among Asian and Black/African American participants—but showed no interaction with income or education. Loneliness showed no race or education interactions with IC, but again a very small interaction with income indicating a slightly weaker association at higher income. A stronger HL–dementia association in Black participants aligns with prior cohort data (ARIC-NCS) showing larger dementia hazards per 10-dB worse hearing among Black than White older adults, and may reflect differences in cumulative vascular and social risks as well as inequities in hearing-care access and device uptake that could amplify downstream cognitive risk (31,32). Given the largely null Loneliness × Education interactions for dementia and only modest attenuation for IC, our results do not provide clear evidence for a cognitive reserve buffering effect (33). Moreover, recent work highlights measurement and cultural/contextual variability in loneliness across racial/ethnic groups, including potential differences in stigma and disclosure and in how loneliness instruments map onto cognitive outcomes—factors that can yield heterogeneous interaction patterns across subgroups (34,35). The apparently stronger loneliness association among those reporting more than one race should be interpreted cautiously given likely small cell sizes and heterogeneity within this category. Overall, these subgroup results suggest that equitable access to hearing care may be especially consequential for cognitive health in some racial/ethnic groups, and that education/cognitive reserve may attenuate the cognitive correlates of loneliness—points that merit confirmation with additional follow-up and repeated psychosocial measures.

### Strengths and Limitations

#### Strengths

The major strengths of this study include its very large sample size, diverse participant pool, longitudinal EHR follow-up, participant responses to surveys of lifestyle and social factors, the usage of time-varying cox analysis, and the ability to adjust for multiple confounders. We also employed a causal mediation analysis, the 4-way decomposition analysis method, to probe mechanisms, adding to the existing literature.

#### Limitations

First, HL status was determined from clinical records, which likely under-ascertains milder cases; without audiometry or onset/duration, severity stratification was not possible, and nondifferential misclassification would bias estimates toward the null. Second, dementia and IC were EHR-ascertained and may be under-recorded, especially with limited access or early disease (36,37); our broader IC codes improve sensitivity but reduce specificity, dementia subtypes were not distinguished, informative censoring (death or loss to follow-up) is possible and competing risk of death was not modeled (the cohort’s younger age may temper this), and missingness in the online SDoH survey could disproportionately exclude cognitively impaired participants, introducing selection bias in loneliness models. Third, loneliness was measured once and HL status was not updated in primary models; both can change over time, potentially attenuating associations, although a time-dependent HL sensitivity analysis suggested minimal short-term impact—longer follow-up with time-varying exposures is warranted. Fourth, residual confounding remains possible despite extensive adjustment (e.g., baseline cognition, social network size, depression, APOE ε4), which is especially relevant for mediation assumptions (no unmeasured mediator–outcome confounding). Finally, hearing-care and loneliness-reduction interventions during follow-up were not modeled and could bias hazard ratios toward the null, and the volunteer nature of AoU participation may limit the generalizability of absolute risks despite the cohort’s diversity.

## CONCLUSION

In this large, diverse AoU cohort, HL and loneliness each predicted higher risk of dementia and IC. The hearing-loss effect appeared to act largely through direct pathways, with a small contribution via loneliness, while loneliness itself independently elevated risk. Looking ahead, further triangulation is needed to strengthen causal inference. Prospective cohorts with repeated measures of hearing, loneliness, and cognition; quasi-experimental studies; Mendelian randomization; and intervention trials targeting hearing rehabilitation and social connection will be important to test whether the patterns observed here reflect modifiable causal pathways. In parallel, health systems and payers can begin to evaluate scalable strategies such as integrating basic hearing screening into routine care, expanding access to affordable hearing devices (including OTC options), and linking patients with HL to resources that support social engagement. Future work with longer follow-up, time-updated measures, and genetic data should clarify mechanisms and identify who benefits most, enabling integrated, life-course approaches to preserve cognitive health.

## Author Contributions

Shuai Yang had full access to all the study data and takes full responsibility for the integrity and accuracy of the reporting.

Conceptualization: Shuai Yang (SY), David A. Sbarra (DAS), Yann C. Klimentidis (YCK)

Methodology: SY, DAS, YCK

Data curation: SY

Software: SY

Formal analysis: SY

Visualization: SY

Writing – original draft: SY

Writing – review & editing: All authors

Supervision: DAS, YCK

## Data Availability

Individual-level electronic health record and survey data were obtained from the NIH AoU Research Program (Controlled Tier v8) and cannot be redistributed outside the secure environment. Qualified researchers may access these data via the All of Us Researcher Workbench after completing registration, required training, and data use agreements. Non-disclosive aggregate outputs underlying the figures and tables are included in the manuscript and Supplement, and analysis code is publicly available (see Code and Materials Availability).

