## Supplementary Material for "Hearing Loss and Loneliness in the Development of Impaired Cognition and Dementia in the All of Us Cohort: Prospective Analyses Using Electronic Health Records"

**Supplementary Table S1. All source diagnosis codes used for hearing loss ascertainment, with OMOP “Maps to” standard SNOMED concepts**

| **#** | **Source vocab** | **Source code** | **Source concept (ICD label)** | **Standard SNOMED concept_id** | **SNOMED code** | **Standard SNOMED concept name** |
| --- | --- | --- | --- | --- | --- | --- |
| 1 | NA | NA | NA | 374053 | 79471008 | Sudden hearing loss |
| 2 | NA | NA | NA | 374366 | 60700002 | Sensorineural hearing loss |
| 3 | NA | NA | NA | 374367 | 95820000 | Bilateral hearing loss |
| 4 | NA | NA | NA | 375826 | 68467004 | Central hearing loss |
| 5 | NA | NA | NA | 376131 | 15190000 | Tympanic membrane conductive hearing loss |
| 6 | NA | NA | NA | 376417 | 61743004 | Middle ear conductive hearing loss |
| 7 | NA | NA | NA | 377574 | 49526009 | Presbycusis |
| 8 | NA | NA | NA | 377888 | 44057004 | Conductive hearing loss |
| 9 | NA | NA | NA | 377889 | 15188001 | Hearing loss |
| 10 | NA | NA | NA | 378442 | 2061000 | Conductive hearing loss of combined sites |
| 11 | NA | NA | NA | 379832 | 77507001 | Mixed conductive AND sensorineural hearing loss |
| 12 | NA | NA | NA | 380734 | 47111006 | External ear conductive hearing loss |
| 13 | NA | NA | NA | 380742 | 3820005 | Inner ear conductive hearing loss |
| 14 | NA | NA | NA | 381312 | 73371001 | Neural hearing loss |
| 15 | NA | NA | NA | 433495 | 194417009 | Conductive hearing loss, bilateral |
| 16 | NA | NA | NA | 436421 | 194429000 | Mixed conductive and sensorineural hearing loss, bilateral |
| 17 | NA | NA | NA | 439709 | 194416000 | Conductive hearing loss due to disorder of inner ear |
| 18 | NA | NA | NA | 439710 | 194415001 | Conductive hearing loss due to disorder of middle ear |
| 19 | NA | NA | NA | 439711 | 194414002 | Conductive hearing loss due to disorder of tympanic membrane |
| 20 | NA | NA | NA | 439712 | 194413008 | Conductive hearing loss due to disorder of external ear |
| 21 | NA | NA | NA | 440422 | 73415002 | Noise-induced hearing loss |
| 22 | NA | NA | NA | 442755 | 65680009 | Sensorineural hearing loss of combined sites |
| 23 | NA | NA | NA | 443577 | 428887009 | Asymmetrical sensorineural hearing loss |
| 24 | NA | NA | NA | 443606 | 430977001 | Bilateral neural hearing loss |
| 25 | NA | NA | NA | 443608 | 430985005 | Bilateral sensory hearing loss |
| 26 | NA | NA | NA | 444291 | 85571008 | Sensory hearing loss |
| 27 | NA | NA | NA | 602624 | 1052205008 | Mixed conductive and sensorineural hearing loss of left ear with normal hearing on right side |
| 28 | NA | NA | NA | 602625 | 1052206009 | Mixed conductive and sensorineural hearing loss of right ear with normal hearing on left side |
| 29 | NA | NA | NA | 605021 | 1010229008 | Neural hearing loss of left ear |
| 30 | NA | NA | NA | 605022 | 1010230003 | Neural hearing loss of right ear |
| 31 | NA | NA | NA | 605027 | 1010236009 | Conductive hearing loss of right ear |
| 32 | NA | NA | NA | 605029 | 1010238005 | Conductive hearing loss of left ear |
| 33 | NA | NA | NA | 605112 | 1010439000 | Conductive hearing loss of left ear with normal hearing on right side |
| 34 | NA | NA | NA | 605113 | 1010440003 | Conductive hearing loss of right ear with normal hearing on left side |
| 35 | NA | NA | NA | 605114 | 1010441004 | Sensorineural hearing loss of right ear with normal hearing on left side |
| 36 | NA | NA | NA | 605115 | 1010442006 | Sensorineural hearing loss of left ear with normal hearing on right side |
| 37 | NA | NA | NA | 605580 | 1119386008 | Sensorineural hearing loss of right ear |
| 38 | NA | NA | NA | 605581 | 1119387004 | Sensorineural hearing loss of left ear |
| 39 | NA | NA | NA | 760154 | 1088891000119106 | Hearing loss in left ear |
| 40 | NA | NA | NA | 761911 | 18701000119107 | Mixed conductive and sensorineural hearing loss of left ear |
| 41 | NA | NA | NA | 761957 | 23631000119109 | Sensorineural hearing loss in left ear |
| 42 | NA | NA | NA | 761958 | 23641000119100 | Sensorineural hearing loss in right ear |
| 43 | NA | NA | NA | 762424 | 3541000119107 | Left conductive hearing loss |
| 44 | NA | NA | NA | 764994 | 1091541000119108 | High-frequency sensorineural hearing loss in right ear |
| 45 | NA | NA | NA | 765124 | 3551000119109 | Right conductive hearing loss |
| 46 | NA | NA | NA | 765313 | 1088931000119103 | High-frequency sensorineural hearing loss in left ear |
| 47 | NA | NA | NA | 765417 | 18711000119105 | Mixed conductive and sensorineural hearing loss of right ear |
| 48 | NA | NA | NA | 4049221 | 232333009 | Hearing loss associated with syndrome |
| 49 | NA | NA | NA | 4107706 | 30169000 | Psychogenic deafness |
| 50 | NA | NA | NA | 4110815 | 194424005 | Sensorineural hearing loss, bilateral |
| 51 | NA | NA | NA | 4146578 | 427644005 | Bilateral central hearing loss |
| 52 | NA | NA | NA | 4176780 | 427772009 | Asymmetrical hearing loss |
| 53 | NA | NA | NA | 36685094 | 1083811000119108 | High-frequency sensorineural hearing loss of bilateral ears |
| 54 | NA | NA | NA | 36685106 | 1084151000119100 | Bilateral hearing loss of ears caused by noise |
| 55 | NA | NA | NA | 36685175 | 1089221000119100 | Hearing loss of left ear caused by noise |
| 56 | NA | NA | NA | 36685246 | 1091831000119101 | Hearing loss of right ear caused by noise |
| 57 | NA | NA | NA | 36715579 | 721294001 | Acquired hearing loss |
| 58 | NA | NA | NA | 37110393 | 724636005 | Sudden idiopathic hearing loss |
| 59 | NA | NA | NA | 37395707 | 715239002 | Sudden sensorineural hearing loss |
| 60 | NA | NA | NA | 40480431 | 441719005 | Speech and language developmental delay due to hearing loss |
| 61 | NA | NA | NA | 42539676 | 737047001 | Mild acquired hearing loss |
| 62 | NA | NA | NA | 43021778 | 473423001 | Hearing loss of right ear |
| 63 | NA | NA | NA | 43021779 | 473424007 | Hearing loss of left ear |
| 64 | NA | NA | NA | 43531411 | 609125008 | Acquired sensorineural hearing loss |
| 65 | NA | NA | NA | 45757561 | 21451000119101 | Mild to moderate hearing loss |
| 66 | NA | NA | NA | 45763917 | 700453005 | Congenital sensorineural hearing loss |
| 67 | NA | NA | NA | 46270115 | 14230001000004101 | Perception of hearing loss |

**Note:** NA indicates that, in the All of Us CDR vocabulary used here, no ICD-9/10 source concept is mapped to this SNOMED standard concept.

**Supplementary Table S2. UCLA Loneliness Scale–8 (ULS-8) items used in the All of Us SDOH survey: stems, response options, and scoring**

| **#** | **Item stem (concise paraphrase)** | **Response options (1–4)** | **Scoring (1–4)** | **Reverse-scored** |
| --- | --- | --- | --- | --- |
| **1** | I lack companionship | Never / Rarely / Sometimes / Often | 1–4 (higher = more lonely) | No |
| **2** | I feel left out | Never / Rarely / Sometimes / Often | 1–4 | No |
| **3** | I feel isolated from others | Never / Rarely / Sometimes / Often | 1–4 | No |
| **4** | I feel close to people | Never / Rarely / Sometimes / Often | Reverse (4→1 … 1→4) | Yes |
| **5** | I feel part of a group of friends | Never / Rarely / Sometimes / Often | Reverse | Yes |
| **6** | There is no one I can turn to | Never / Rarely / Sometimes / Often | 1–4 | No |
| **7** | I am outgoing / sociable | Never / Rarely / Sometimes / Often | Reverse | Yes |
| **8** | People are around me but not with me | Never / Rarely / Sometimes / Often | 1–4 | No |

**Supplementary Table S3. All source diagnosis codes used for dementia ascertainment, with OMOP “Maps to” standard SNOMED concepts**

| **#** | **Source vocab** | **Source code** | **Source concept (ICD label)** | **Standard SNOMED concept_id** | **SNOMED code** | **Standard SNOMED concept name** |
| --- | --- | --- | --- | --- | --- | --- |
| 1 | ICD10CM | F06.0 | Psychotic disorder with hallucinations due to known physiological condition | 373175 | 45912004 | Organic hallucinosis |
| 2 | ICD10CM | F06.8 | Other specified mental disorders due to known physiological condition | 374009 | 111479008 | Organic mental disorder |
| 3 | ICD10CM | G30.9 | Alzheimer’s disease, unspecified | 378419 | 26929004 | Alzheimer’s disease |
| 4 | ICD9CM | 290.21 | Senile dementia with depressive features | 379784 | 191459006 | Senile dementia with depression |
| 5 | ICD9CM | 291.2 | Alcohol-induced persisting dementia | 378726 | 281004 | Dementia associated with alcoholism |
| 6 | ICD9CM | 290.20 | Senile dementia with delusional features | 380986 | 371024007 | Senile dementia with delusion |
| 7 | ICD9CM | 290.11 | Presenile dementia with delirium | 381832 | 191452002 | Presenile dementia with delirium |
| 8 | ICD9CM | 290.42 | Vascular dementia, with delusions | 443790 | 25772007 | Multi-infarct dementia with delusions |
| 9 | ICD9CM | 290.41 | Vascular dementia, with delirium | 444091 | 10349009 | Multi-infarct dementia with delirium |
| 10 | ICD9CM | 291.1 | Alcohol-induced persisting amnestic disorder | 374623 | 73097000 | Alcohol amnestic disorder |
| 11 | ICD9CM | 291.0 | Alcohol withdrawal delirium | 377830 | 8635005 | Alcohol withdrawal delirium |
| 12 | ICD9CM | 292.82 | Drug-induced persisting dementia | 376095 | 191493005 | Drug-induced dementia |
| 13 | ICD9CM | 331.0 | Alzheimer’s disease | 378419 | 26929004 | Alzheimer’s disease |
| 14 | ICD9CM | 290.40 | Vascular dementia, uncomplicated | 377254 | 70936005 | Multi-infarct dementia, uncomplicated |
| 15 | ICD9CM | 294.11 | Dementia in conditions classified elsewhere with behavioral disturbance | 43530666 | 1591000119103 | Dementia with behavioral disturbance |
| 16 | ICD9CM | 294.11 | Dementia in conditions classified elsewhere with behavioral disturbance | 374888 | 191519005 | Dementia associated with another disease |
| 17 | ICD9CM | 294.8 | Other persistent mental disorders due to conditions classified elsewhere | 374009 | 111479008 | Organic mental disorder |
| 18 | ICD9CM | 290.43 | Vascular dementia, with depressed mood | 443864 | 14070001 | Multi-infarct dementia with depression |
| 19 | ICD9CM | 294.10 | Dementia in conditions classified elsewhere without behavioral disturbance | 374888 | 191519005 | Dementia associated with another disease |
| 20 | ICD9CM | 290.3 | Senile dementia with delirium | 376946 | 191461002 | Senile dementia with delirium |
| 21 | ICD9CM | 331.82 | Dementia with lewy bodies | 380701 | 80098002 | Diffuse Lewy body disease |
| 22 | ICD9CM | 290.12 | Presenile dementia with delusions | 44782771 | 31081000119101 | Presenile dementia with delusions |
| 23 | ICD9CM | 290.0 | Senile dementia, uncomplicated | 375791 | 191449005 | Uncomplicated senile dementia |
| 24 | ICD9CM | 290.10 | Presenile dementia, uncomplicated | 376085 | 191451009 | Uncomplicated presenile dementia |
| 25 | ICD9CM | 331.11 | Pick’s disease | 4043378 | 230270009 | Frontotemporal dementia |
| 26 | ICD9CM | 290.13 | Presenile dementia with depressive features | 377527 | 191455000 | Presenile dementia with depression |
| 27 | ICD10CM | F03.90 | Unspecified dementia, unspecified severity, without behavioral disturbance… | 4182210 | 52448006 | Dementia |
| 28 | ICD10CM | G31.01 | Frontotemporal dementia | 4043378 | 230270009 | Frontotemporal dementia |
| 29 | ICD10CM | F19.97 | Other psychoactive substance use, unspecified with … persisting dementia | 4009647 | 111480006 | Psychoactive substance-induced organic dementia |
| 30 | ICD10CM | G31.83 | Neurocognitive disorder with Lewy bodies | 4196433 | 312991009 | Senile dementia of the Lewy body type |
| 31 | ICD10CM | F10.231 | Alcohol dependence with withdrawal delirium | 435243 | 66590003 | Alcohol dependence |
| 32 | ICD10CM | F10.231 | Alcohol dependence with withdrawal delirium | 377830 | 8635005 | Alcohol withdrawal delirium |
| 33 | ICD10CM | F10.27 | Alcohol use, unspecified with alcohol-induced persisting dementia | 435243 | 66590003 | Alcohol dependence |
| 34 | ICD10CM | F10.27 | Alcohol use, unspecified with alcohol-induced persisting dementia | 378726 | 281004 | Dementia associated with alcoholism |
| 35 | ICD10CM | F10.96 | Alcohol use, unspecified with alcohol-induced persisting amnestic disorder | 374623 | 73097000 | Alcohol amnestic disorder |
| 36 | ICD10CM | F01.50 | Vascular dementia, unspecified severity, without behavioral disturbance… | 37109056 | 16276361000119109 | Vascular dementia without behavioral disturbance |
| 37 | ICD10CM | F02.80 | Dementia in other diseases classified elsewhere, w/o behavioral disturbance | 374888 | 191519005 | Dementia associated with another disease |
| 38 | ICD10CM | F02.81 | Dementia in other diseases classified elsewhere, with behavioral disturbance | 43530666 | 1591000119103 | Dementia with behavioral disturbance |
| 39 | ICD10CM | F02.81 | Dementia in other diseases classified elsewhere, with behavioral disturbance | 374888 | 191519005 | Dementia associated with another disease |
| 40 | ICD10CM | F03.91 | Unspecified dementia, unspecified severity, with behavioral disturbance | 43530666 | 1591000119103 | Dementia with behavioral disturbance |
| 41 | ICD10CM | F01.51 | Vascular dementia, unspecified severity, with behavioral disturbance | 37018688 | 288631000119104 | Vascular dementia with behavioral disturbance |

**Supplementary Table S4. All source diagnosis codes used for Impaired cognition ascertainment, with OMOP “Maps to” standard SNOMED concepts**

| **#** | **Source vocab** | **Source code** | **Source concept (ICD label)** | **Standard SNOMED concept_id** | **SNOMED code** | **Standard SNOMED concept name** |
| --- | --- | --- | --- | --- | --- | --- |
| 1 | NA | NA | NA | 437306 | 230736007 | Transient global amnesia |
| 2 | NA | NA | NA | 439147 | 48167000 | Amnesia |
| 3 | NA | NA | NA | 439795 | 110352000 | Minimal cognitive impairment |
| 4 | NA | NA | NA | 443432 | 386806002 | Impaired cognition |
| 5 | NA | NA | NA | 4009705 | 102891000 | Age-related cognitive decline |
| 6 | NA | NA | NA | 4012209 | 162200009 | Temporary loss of memory |
| 7 | NA | NA | NA | 4074319 | 225037001 | Minor memory lapses |
| 8 | NA | NA | NA | 4076654 | 225038006 | Memory lapses |
| 9 | NA | NA | NA | 4099961 | 192071009 | Mild memory disturbance |
| 10 | NA | NA | NA | 4145069 | 307413004 | Transient memory loss |
| 11 | NA | NA | NA | 4198081 | 51921000 | Retrograde amnesia |
| 12 | NA | NA | NA | 4206332 | 55533009 | Forgetful |
| 13 | NA | NA | NA | 4229448 | 88822006 | Anterograde amnesia |
| 14 | NA | NA | NA | 4304008 | 386807006 | Memory impairment |
| 15 | NA | NA | NA | 36687122 | 15928141000119107 | Human immunodeficiency virus infection with cognitive impairment |
| 16 | NA | NA | NA | 37109222 | 2421000119107 | Hallucinations co-occurrent and due to late onset dementia |
| 17 | NA | NA | NA | 37117145 | 16219201000119101 | Behavioral disturbance co-occurrent and due to late onset Alzheimer dementia |
| 18 | NA | NA | NA | 42537141 | 736317001 | Impaired executive functioning |
| 19 | NA | NA | NA | 44782432 | 105421000119105 | Early onset Alzheimer’s disease with behavioral disturbance |
| 20 | NA | NA | NA | 44782725 | 141601000119107 | Cognitive changes due to organic disorder |
| 21 | NA | NA | NA | 44784521 | 698687007 | Post-traumatic dementia with behavioral change |
| 22 | NA | NA | NA | 44784643 | 97751000119108 | Altered behavior in Alzheimer’s disease |
| 23 | NA | NA | NA | 45765899 | 702955000 | Moderate cognitive impairment |
| 24 | NA | NA | NA | 45765900 | 702956004 | Severe cognitive impairment |

**Supplementary Table S5. Multiple linear regression of loneliness (ULS-8 mean, item-average) on baseline hearing loss and covariates (N = 101,525)
*(Unstandardized coefficients; ULS-8 mean × 8 = ULS-8 total)*)**

| **Variable** | **β (Estimate)** | **SE** | **95% CI** | **p-value** |
| --- | --- | --- | --- | --- |
| **Hearing loss (yes vs no)** | 0.035 | 0.005 | 0.025, 0.045 | <0.001 |
| **Age (per year)** | −0.0067 | 0.0001 | −0.0069, −0.0064 | <0.001 |
| **Gender** |  |  |  |  |
| **Male vs Female** | 0.011 | 0.004 | 0.003, 0.019 | 0.006 |
| **Non-binary vs Female** | 0.319 | 0.028 | 0.263, 0.375 | <0.001 |
| **Race/Ethnicity (ref: White)** |  |  |  |  |
| **American Indian or Alaska Native** | −0.048 | 0.026 | −0.098, 0.003 | 0.063 |
| **Asian** | 0.032 | 0.012 | 0.008, 0.056 | 0.010 |
| **Black or African American** | −0.050 | 0.007 | −0.064, −0.036 | <0.001 |
| **Middle Eastern or North African** | 0.033 | 0.028 | −0.022, 0.089 | 0.233 |
| **More than one population** | 0.077 | 0.010 | 0.058, 0.096 | <0.001 |
| **Native Hawaiian or Other Pacific Islander** | −0.192 | 0.097 | −0.383, −0.001 | 0.049 |
| **Education (per year)** | 0.0033 | 0.0011 | 0.0012, 0.0054 | 0.002 |
| **Income (per dollar)** | −2.427×10⁻⁶ | 3.295×10⁻⁸ | −2.492×10⁻⁶, −2.362×10⁻⁶ | <0.001 |
| **Smoking (ever/current vs never)** | 0.053 | 0.004 | 0.045, 0.061 | <0.001 |
| **Drinking (ever/current vs never)** | 0.003 | 0.011 | −0.018, 0.024 | 0.768 |

**Notes:**
ULS-8 = UCLA Loneliness Scale–8 item version.
β = regression coefficient; SE = standard error; CI = confidence interval.
Model adjusted for all variables in table.

**Supplementary Table S6. Associations Between Hearing Loss and Dementia, With Interaction by Race, Income, and Education**

| **Variable** | **HR** | **95% CI** | **p-value** |
| --- | --- | --- | --- |
| **Model 1: Hearing Loss × Race** |  |  |  |
| Hearing loss | 1.19 | 1.10–1.29 | <0.001 |
| American Indian or Alaska Native | 2.15 | 1.61–2.87 | <0.001 |
| Asian | 1.01 | 0.72–1.42 | 0.943 |
| Black or African American | 0.80 | 0.71–0.90 | <0.001 |
| Middle Eastern or North African | 0.78 | 0.37–1.64 | 0.509 |
| More than one population | 1.41 | 1.17–1.69 | <0.001 |
| Native Hawaiian or Other Pacific Islander | —* | —* | 0.974 |
| Age at first visit (per year) | 1.06 | 1.056–1.062 | <0.001 |
| Male (vs female) | 1.52 | 1.42–1.62 | <0.001 |
| Non-binary (vs female) | 2.45 | 1.10–5.46 | 0.029 |
| Education (per year) | 0.96 | 0.95–0.98 | <0.001 |
| Income (per $1) | 1.00 | 1.00–1.00 | <0.001 |
| Current smoking | 1.16 | 1.08–1.24 | <0.001 |
| Current drinking | 0.71 | 0.62–0.81 | <0.001 |
| HL × American Indian or Alaska Native | 0.82 | 0.46–1.48 | 0.516 |
| HL × Asian | 1.21 | 0.66–2.19 | 0.536 |
| HL × Black or African American | 1.76 | 1.45–2.13 | <0.001 |
| HL × Middle Eastern or North African | 1.32 | 0.42–4.17 | 0.637 |
| HL × More than one population | 1.09 | 0.80–1.48 | 0.581 |
| HL × Native Hawaiian or Other Pacific Islander | 0.55 | —* | 0.999 |
| **Model 2: Hearing Loss × Income** |  |  |  |
| Hearing loss (HL) | 1.18 | 1.06–1.32 | 0.003 |
| Income (per $1) | 1.00 | 1.00–1.00 | <0.001 |
| Age at first visit (per year) | 1.06 | 1.056–1.062 | <0.001 |
| Male (vs female) | 1.49 | 1.39–1.60 | <0.001 |
| Non-binary (vs female) | 2.49 | 1.12–5.56 | 0.026 |
| American Indian or Alaska Native | 2.07 | 1.61–2.67 | <0.001 |
| Asian | 1.08 | 0.82–1.43 | 0.572 |
| Black or African American | 0.93 | 0.85–1.03 | 0.171 |
| Middle Eastern or North African | 0.87 | 0.50–1.54 | 0.642 |
| More than one population | 1.46 | 1.26–1.69 | <0.001 |
| Native Hawaiian or Other Pacific Islander | —* | —* | 0.970 |
| Education (per year) | 0.96 | 0.95–0.98 | <0.001 |
| Current smoking | 1.15 | 1.07–1.23 | <0.001 |
| Current drinking | 0.71 | 0.62–0.81 | <0.001 |
| HL × Income | 1.00 | 1.00–1.00 | 0.030 |
| **Model 3: Hearing Loss × Education** |  |  |  |
| Hearing loss (HL) | 1.01 | 0.67–1.54 | 0.950 |
| Education (per year) | 0.96 | 0.94–0.98 | <0.001 |
| Age at first visit (per year) | 1.06 | 1.056–1.062 | <0.001 |
| Male (vs female) | 1.49 | 1.40–1.60 | <0.001 |
| Non-binary (vs female) | 2.49 | 1.12–5.56 | 0.026 |
| American Indian or Alaska Native | 2.07 | 1.61–2.67 | <0.001 |
| Asian | 1.08 | 0.82–1.43 | 0.575 |
| Black or African American | 0.94 | 0.85–1.04 | 0.204 |
| Middle Eastern or North African | 0.88 | 0.50–1.55 | 0.645 |
| More than one population | 1.46 | 1.26–1.70 | <0.001 |
| Native Hawaiian or Other Pacific Islander | —* | —* | 0.970 |
| Income (per $1) | 1.00 | 1.00–1.00 | <0.001 |
| Current smoking | 1.15 | 1.08–1.24 | <0.001 |
| Current drinking | 0.71 | 0.62–0.81 | <0.001 |
| HL × Education (per year) | 1.02 | 0.99–1.05 | 0.245 |

**Supplementary Table S7. Associations Between Loneliness and Dementia, With Interaction by Race, Income, and Education**

| **Variable** | **HR** | **95% CI** | **p-value** |
| --- | --- | --- | --- |
| **Model 1: Loneliness × Race** |  |  |  |
| Loneliness score (per unit) | 1.71 | 1.56–1.89 | <0.001 |
| American Indian or Alaska Native | 4.26 | 0.60–30.04 | 0.146 |
| Asian | 0.83 | 0.16–4.44 | 0.828 |
| Black or African American | 1.51 | 0.76–3.00 | 0.236 |
| Middle Eastern or North African | 0.06 | 0.00–12.67 | 0.309 |
| More than one population | 2.54 | 1.01–6.41 | 0.048 |
| Native Hawaiian or Other Pacific Islander | —* | —* | 0.993 |
| Age at loneliness (per year) | 1.05 | 1.05–1.06 | <0.001 |
| Male (vs female) | 1.56 | 1.38–1.76 | <0.001 |
| Non-binary (vs female) | 2.59 | 0.96–6.96 | 0.059 |
| Education (per year) | 0.98 | 0.95–1.01 | 0.128 |
| Income (per $1) | 1.00 | 1.00–1.00 | <0.001 |
| Current smoking | 1.09 | 0.96–1.23 | 0.186 |
| Current drinking | 0.54 | 0.41–0.71 | <0.001 |
| Loneliness × American Indian/Alaska Native | 0.65 | 0.26–1.63 | 0.357 |
| Loneliness × Asian | 1.11 | 0.53–2.34 | 0.785 |
| Loneliness × Black/African American | 0.81 | 0.60–1.10 | 0.182 |
| Loneliness × Middle Eastern/North African | 2.33 | 0.33–16.23 | 0.394 |
| Loneliness × More than one population | 0.74 | 0.49–1.12 | 0.151 |
| Loneliness × Native Hawaiian/Other Pacific Islander | —* | —* | 1.000 |
| **Model 2: Loneliness × Income** |  |  |  |
| Loneliness score (per unit) | 1.87 | 1.63–2.15 | <0.001 |
| Income (per $1) | 1.00 | 1.00–1.00 | 0.768 |
| Age at loneliness (per year) | 1.05 | 1.05–1.06 | <0.001 |
| Male (vs female) | 1.56 | 1.38–1.76 | <0.001 |
| Non-binary (vs female) | 2.53 | 0.94–6.79 | 0.066 |
| American Indian or Alaska Native | 1.70 | 0.91–3.19 | 0.095 |
| Asian | 1.04 | 0.62–1.74 | 0.887 |
| Black or African American | 0.96 | 0.77–1.20 | 0.733 |
| Middle Eastern or North African | 0.49 | 0.12–1.97 | 0.317 |
| More than one population | 1.30 | 0.98–1.74 | 0.073 |
| Native Hawaiian or Other Pacific Islander | —* | —* | 0.980 |
| Education (per year) | 0.98 | 0.95–1.01 | 0.159 |
| Current smoking | 1.08 | 0.96–1.22 | 0.203 |
| Current drinking | 0.54 | 0.41–0.71 | <0.001 |
| Loneliness × Income | 1.00 | 1.00–1.00 | 0.030 |
| **Model 3: Loneliness × Education** |  |  |  |
| Loneliness score (per unit) | 2.07 | 1.13–3.80 | 0.019 |
| Education (per year) | 1.01 | 0.92–1.10 | 0.870 |
| Age at loneliness (per year) | 1.05 | 1.05–1.06 | <0.001 |
| Male (vs female) | 1.56 | 1.38–1.76 | <0.001 |
| Non-binary (vs female) | 2.58 | 0.96–6.93 | 0.060 |
| American Indian or Alaska Native | 1.71 | 0.91–3.20 | 0.093 |
| Asian | 1.04 | 0.62–1.73 | 0.896 |
| Black or African American | 0.96 | 0.77–1.21 | 0.751 |
| Middle Eastern or North African | 0.49 | 0.12–1.97 | 0.315 |
| More than one population | 1.31 | 0.98–1.75 | 0.068 |
| Native Hawaiian or Other Pacific Islander | —* | —* | 0.980 |
| Income (per $1) | 1.00 | 1.00–1.00 | <0.001 |
| Current smoking | 1.09 | 0.96–1.23 | 0.193 |
| Current drinking | 0.54 | 0.41–0.71 | <0.001 |
| Loneliness × Education (per year) | 0.99 | 0.95–1.03 | 0.466 |

**Supplementary Table S8. Associations Between Hearing Loss and Impaired Cognition, With Interaction by Race, Income, and Education**

| **Variable** | **HR** | **95% CI** | **p-value** | |
| --- | --- | --- | --- | --- |
| **Model 1: Hearing Loss × Race** |  |  | |  |
| Hearing loss (HL) | 1.56 | 1.50–1.62 | | <0.001 |
| American Indian/Alaska Native | 1.00 | 0.80–1.25 | | 0.983 |
| Asian | 0.87 | 0.74–1.03 | | 0.106 |
| Black or African American | 0.66 | 0.62–0.71 | | <0.001 |
| Middle Eastern or North African | 1.26 | 0.95–1.69 | | 0.109 |
| More than one population | 1.28 | 1.16–1.41 | | <0.001 |
| Native Hawaiian or Other Pacific Islander | 0.89 | 0.34–2.38 | | 0.822 |
| White | Reference | — | | — |
| Age (per year) | 1.04 | 1.04–1.04 | | <0.001 |
| Male (vs female) | 0.87 | 0.84–0.91 | | <0.001 |
| Non-binary (vs female) | 2.28 | 1.65–3.14 | | <0.001 |
| Education (per year) | 1.00 | 0.99–1.01 | | 0.784 |
| Income (per $1) | 1.00 | 1.00–1.00 | | <0.001 |
| Current smoking | 1.00 | 0.97–1.04 | | 0.989 |
| Current drinking | 0.93 | 0.87–1.01 | | 0.083 |
| HL × American Indian/Alaska Native | 1.24 | 0.85–1.80 | | 0.267 |
| HL × Asian | 1.42 | 1.08–1.87 | | 0.013 |
| HL × Black or African American | 1.52 | 1.37–1.69 | | <0.001 |
| HL × Middle Eastern or North African | 0.97 | 0.60–1.57 | | 0.905 |
| HL × More than one population | 1.08 | 0.93–1.26 | | 0.361 |
| HL × Native Hawaiian or Other Pacific Islander | 1.46 | 0.33–6.51 | | 0.623 |
| **Model 2: Hearing Loss × Income** |  |  | |  |
| **Variable** | **HR** | **95% CI** | | **p-value** |
| Hearing loss (HL) | **1.63** | **1.55–1.73** | | **<0.001** |
| Income (per $1) | **1.00** | **1.00–1.00** | | **<0.001** |
| Age (per year) | **1.04** | **1.04–1.04** | | **<0.001** |
| Male (vs female) | **0.87** | **0.84–0.90** | | **<0.001** |
| Non-binary (vs female) | **2.30** | **1.67–3.16** | | **<0.001** |
| American Indian/Alaska Native | 1.09 | 0.91–1.30 | | 0.372 |
| Asian | 0.98 | 0.86–1.12 | | 0.800 |
| Black or African American | **0.75** | **0.71–0.79** | | **<0.001** |
| Middle Eastern or North African | 1.26 | 1.00–1.58 | | 0.051 |
| More than one population | **1.32** | **1.22–1.42** | | **<0.001** |
| Native Hawaiian or Other Pacific Islander | 1.04 | 0.50–2.19 | | 0.914 |
| White | **Reference** | — | | — |
| Education (per year) | 1.00 | 0.99–1.01 | | 0.916 |
| Current smoking | 1.00 | 0.96–1.03 | | 0.915 |
| Current drinking | 0.94 | 0.87–1.01 | | 0.088 |
| HL × Income | 1.00 | 1.00–1.00 | | 0.592 |
| **Model 3: Hearing Loss × Education** |  |  | |  |
| **Variable** | **HR** | **95% CI** | | **p-value** |
| Hearing loss (HL) | 1.88 | 1.51–2.35 | | <0.001 |
| Education (per year) | 1.00 | 0.99–1.01 | | 0.551 |
| Age (per year) | 1.04 | 1.04–1.04 | | <0.001 |
| Male (vs female) | 0.87 | 0.84–0.90 | | <0.001 |
| Non-binary (vs female) | 2.30 | 1.67–3.16 | | <0.001 |
| American Indian/Alaska Native | 1.09 | 0.91–1.31 | | 0.350 |
| Asian | 0.98 | 0.86–1.12 | | 0.788 |
| Black or African American | 0.76 | 0.72–0.80 | | <0.001 |
| Middle Eastern or North African | 1.26 | 1.00–1.58 | | 0.053 |
| More than one population | 1.32 | 1.22–1.43 | | <0.001 |
| Native Hawaiian or Other Pacific Islander | 1.04 | 0.50–2.19 | | 0.911 |
| White | Reference | — | | — |
| Income (per $1) | 1.00 | 1.00–1.00 | | <0.001 |
| Current smoking | 1.00 | 0.97–1.03 | | 0.953 |
| Current drinking | 0.93 | 0.87–1.01 | | 0.084 |
| HL × Education (per year) | 0.99 | 0.98–1.01 | | 0.242 |

**Supplementary Table S9. Associations Between Loneliness and Impaired Cognition, With Interaction by Race, Income, and Education**

| **Variable** | **HR** | **95% CI** | **p-value** |
| --- | --- | --- | --- |
| **Model 1: Loneliness × Race** |  |  |  |
| Loneliness (ULS-8 mean, per 1-point) | 1.58 | 1.52–1.64 | <0.001 |
| American Indian/Alaska Native | 0.96 | 0.34–2.72 | 0.940 |
| Asian | 1.11 | 0.57–2.17 | 0.767 |
| Black/African American | 0.99 | 0.74–1.32 | 0.926 |
| Middle Eastern/North African | 1.74 | 0.60–5.05 | 0.310 |
| More than one population | 0.99 | 0.67–1.45 | 0.952 |
| Native Hawaiian/Other Pacific Islander | 2.57 | 0.09–70.96 | 0.576 |
| Age (per year) | 1.04 | 1.04–1.04 | <0.001 |
| Male (vs female) | 0.94 | 0.90–0.99 | 0.020 |
| Non-binary (vs female) | 2.13 | 1.47–3.09 | <0.001 |
| Education (per year) | 0.98 | 0.97–0.99 | 0.001 |
| Income (per $1) | 1.00 | 1.00–1.00 | <0.001 |
| Current smoking | 1.02 | 0.98–1.08 | 0.328 |
| Current drinking | 0.87 | 0.77–1.00 | 0.045 |
| Loneliness × American Indian/Alaska Native | 1.09 | 0.69–1.70 | 0.712 |
| Loneliness × Asian | 0.92 | 0.67–1.26 | 0.599 |
| Loneliness × Black/African American | 0.91 | 0.79–1.03 | 0.139 |
| Loneliness × Middle Eastern/North African | 0.82 | 0.50–1.35 | 0.436 |
| Loneliness × More than one population | 1.11 | 0.95–1.31 | 0.196 |
| Loneliness × Native Hawaiian/Other Pacific Islander | 0.73 | 0.12–4.35 | 0.732 |
| **Model 2: Loneliness × Income** |  |  |  |
| Loneliness (ULS-8 mean, per 1-point) | 1.65 | 1.56–1.75 | <0.001 |
| Income (per $1) | 1.00 | 1.00–1.00 | 0.051 |
| Age (per year) | 1.04 | 1.04–1.04 | <0.001 |
| Male (vs female) | 0.94 | 0.90–0.99 | 0.018 |
| Non-binary (vs female) | 2.12 | 1.46–3.08 | <0.001 |
| American Indian/Alaska Native | 1.15 | 0.84–1.58 | 0.368 |
| Asian | 0.93 | 0.76–1.15 | 0.515 |
| Black/African American | 0.80 | 0.73–0.88 | <0.001 |
| Middle Eastern/North African | 1.16 | 0.80–1.67 | 0.430 |
| More than one population | 1.26 | 1.12–1.41 | <0.001 |
| Native Hawaiian/Other Pacific Islander | 1.47 | 0.47–4.56 | 0.505 |
| Education (per year) | 0.98 | 0.97–0.99 | 0.002 |
| Current smoking | 1.02 | 0.97–1.07 | 0.354 |
| Current drinking | 0.87 | 0.77–1.00 | 0.045 |
| Loneliness × Income | 1.00 | 1.00–1.00 | 0.033 |
| **Model 3: Loneliness × Education** |  |  |  |
| Loneliness (ULS-8 mean, per 1-point) | 1.77 | 1.38–2.26 | <0.001 |
| Education (per year) | 1.00 | 0.96–1.03 | 0.808 |
| Age (per year) | 1.04 | 1.04–1.04 | <0.001 |
| Male (vs female) | 0.94 | 0.90–0.99 | 0.019 |
| Non-binary (vs female) | 2.13 | 1.47–3.09 | <0.001 |
| American Indian/Alaska Native | 1.15 | 0.85–1.58 | 0.367 |
| Asian | 0.93 | 0.76–1.15 | 0.508 |
| Black/African American | 0.80 | 0.73–0.88 | <0.001 |
| Middle Eastern/North African | 1.16 | 0.80–1.67 | 0.433 |
| More than one population | 1.26 | 1.12–1.41 | <0.001 |
| Native Hawaiian/Other Pacific Islander | 1.47 | 0.47–4.55 | 0.507 |
| Income (per $1) | 1.00 | 1.00–1.00 | <0.001 |
| Current smoking | 1.02 | 0.98–1.07 | 0.342 |
| Current drinking | 0.87 | 0.77–1.00 | 0.045 |
| Loneliness × Education (per year) | 0.99 | 0.98–1.01 | 0.355 |

**Supplementary Table S10. Time-varying Cox proportional hazards model of hearing loss and dementia, adjusted for demographics, socioeconomic factors, and health behaviors (n= 241695, number of events= 4319)**

| **Variable** | **HR** | **95% CI** | **p-value** |
| --- | --- | --- | --- |
| Hearing loss | 1.10 | 1.02–1.19 | 0.018 |
| Age (per year) | 1.06 | 1.06–1.06 | <0.001 |
| Male (vs Female) | 1.57 | 1.47–1.67 | <0.001 |
| Non-Binary (vs Female) | 2.40 | 1.14–5.04 | 0.021 |
| American Indian or Alaska Native | 1.85 | 1.45–2.35 | <0.001 |
| Asian | 1.01 | 0.79–1.30 | 0.936 |
| Black or African American | 0.86 | 0.79–0.94 | 0.001 |
| Middle Eastern or North African | 0.79 | 0.47–1.34 | 0.387 |
| More than one population | 1.37 | 1.20–1.58 | <0.001 |
| Native Hawaiian or Other Pacific Islander | ~0 | — | 0.966 |
| Education (years) | 0.98 | 0.96–0.99 | 0.001 |
| Income (per $1) | 1.00 | 1.00–1.00 | <0.001 |
| Smoking | 1.13 | 1.06–1.21 | <0.001 |
| Drinking | 0.70 | 0.62–0.79 | <0.001 |

**Supplementary Table S11. Time-varying Cox proportional hazards model for the association between hearing loss and impaired cognition (n= 239648, number of events= 17643)**

| **Variable** | **HR** | **95% CI** | **p-value** |
| --- | --- | --- | --- |
| Hearing loss | 1.40 | 1.36–1.44 | <0.001 |
| Age (per year) | 1.04 | 1.04–1.04 | <0.001 |
| Male (vs Female) | 0.97 | 0.94–0.99 | 0.023 |
| Non-Binary (vs Female) | 2.20 | 1.65–2.94 | <0.001 |
| American Indian or Alaska Native | 1.00 | 0.84–1.19 | 0.913 |
| Asian | 0.91 | 0.80–1.03 | 0.143 |
| Black or African American | 0.69 | 0.65–0.74 | <0.001 |
| Middle Eastern or North African | 1.19 | 0.96–1.48 | 0.100 |
| More than one population | 1.28 | 1.17–1.40 | <0.001 |
| Native Hawaiian or Other Pacific Islander | 1.01 | 0.53–1.91 | 0.984 |
| Education (years) | 1.01 | 1.00–1.02 | 0.055 |
| Income (per $1) | 1.00 | 1.00–1.00 | <0.001 |
| Smoking | 1.02 | 0.99–1.06 | 0.334 |
| Drinking | 0.94 | 0.88–1.00 | 0.070 |

**Supplementary Methods S1. Time-varying Cox sensitivity analysis**

We implemented a time-dependent Cox model in counting-process (“long”) format to allow hearing loss (HL) status to change over follow-up. Each participant contributed an interval with HL=0 until the first EHR code for HL and, if applicable, subsequent intervals with HL=1. The outcome (dementia or impaired cognition) was modeled using Cox proportional hazards with the same covariate set as in the primary analyses; standard errors were clustered at the participant level. Loneliness was not time-updated due to the absence of repeated measures and was retained as a baseline variable. This approach aligns with multi-state/time-dependent covariate methods and helps mitigate exposure misclassification from incident HL after baseline.

**Supplementary Results SR1.**

Relative to baseline-exposure models, time-varying HL models yielded very similar associations: for dementia, the time-varying HL hazard ratio (HR) was 1.10 (95% CI: 1.02–1.19, p = 0.018); for impaired cognition, the HR was 1.40 (95% CI: 1.36–1.44, p < 0.001). These results indicate minimal sensitivity to treating HL as time-varying and suggest limited bias from baseline exposure misclassification.
